# Improving genetic risk modeling of dementia from real-world data in underrepresented populations

**DOI:** 10.1101/2024.02.05.24302355

**Authors:** Mingzhou Fu, Leopoldo Valiente-Banuet, Satpal S. Wadhwa, UCLA Precision Health Data Discovery Repository Working Group, UCLA Precision Health ATLAS Working Group, Bogdan Pasaniuc, Keith Vossel, Timothy S. Chang

## Abstract

**BACKGROUND:** Genetic risk modeling for dementia offers significant benefits, but studies based on real-world data, particularly for underrepresented populations, are limited.

**METHODS:** We employed an Elastic Net model for dementia risk prediction using single-nucleotide polymorphisms prioritized by functional genomic data from multiple neurodegenerative disease genome-wide association studies. We compared this model with *APOE* and polygenic risk score models across genetic ancestry groups, using electronic health records from UCLA Health for discovery and All of Us cohort for validation.

**RESULTS:** Our model significantly outperforms other models across multiple ancestries, improving the area-under-precision-recall curve by 21-61% and the area-under-the-receiver-operating characteristic by 10-21% compared to the *APOE* and the polygenic risk score models. We identified shared and ancestry-specific risk genes and biological pathways, reinforcing and adding to existing knowledge.

**CONCLUSIONS:** Our study highlights benefits of integrating functional mapping, multiple neurodegenerative diseases, and machine learning for genetic risk models in diverse populations. Our findings hold potential for refining precision medicine strategies in dementia diagnosis.

## 1 Background

Dementia, a complex and multifaceted syndrome, is characterized by a progressive decline in cognitive function beyond what might be expected from normal aging. Etiologies include Alzheimer’s disease (AD), vascular dementia, Lewy body dementia (LBD), Frontotemporal dementia (FTD), and Parkinson’s disease dementia (PDD), among others.^1^ The prognosis of dementia is generally a gradual and continuous decline in cognitive function, which can significantly impact an individual’s ability to perform daily activities.^2^ Dementia represents a significant public health concern, with a global prevalence estimated at around 36 million in 2020. Owing to an aging population, this number is projected to triple by 2050.^3^ The economic burden of dementia is also substantial, with global costs estimated to be around $594 billion annually.^4^

Dementia has a strong genetic predisposition, with numerous significant genetic variants associated with the disease identified through Genome-Wide Association Studies (GWASs). For example, the Apolipoprotein E (*APOE)* gene, which encodes a protein responsible for binding and transporting low-density lipids, significantly influences the risk of late-onset AD, the most prevalent form of dementia.^5,6^ Similarly, the Microtubule-associated protein tau (*MAPT*) is a recognized genetic mutation in FTD,^7^ and Synuclein Alpha (*SNCA*) is associated with PDD.^8^ While these studies have deepened our understanding of the genetic architecture of dementia, additional research is necessary to successfully model personal dementia genetic risk and understand the potential limitations.

Polygenic risk scores (PRSs), which aggregate the effects of many genetic variants associated with a disease, have recently been used to quantify an individual’s genetic predisposition for complex diseases like dementia.^9^ A growing number of studies have underscored the robust links between AD PRS and AD phenotype,^10–13^ declines in memory and executive function,^14–17^ clinical progression,^15^ and amyloid load^18^ in the non-Hispanic white population. However, the performance of PRSs in non-European ancestries has been suboptimal. The weights for SNPs in PRSs are predominantly calculated based on European ancestry GWASs, leading to a lack of generalizability in representing genetic risks for non-European individuals.^19–22^ Using PRSs for 245 curated traits from the UK Biobank data, Privé et al.^23^ revealed notable disparities in the phenotypic variance explained by PRSs across different populations. Specifically, compared to individuals of Northwestern European ancestry, the PRS-driven phenotypic variance is only 64.7% in South Asians, 48.6% in East Asians, and 18% in West Africans. Similarly, using a population from the Health and Retirement Study, Marden et al. demonstrated that the estimated effect of the AD PRS was notably smaller for non-Hispanic black compared to non-Hispanic white in both dementia probability score and memory score.^24^

Another limitation of current genetic risk modeling is differentiating between causal and uninformative variants. Causal variants, such as *APOE* in AD, have been suggested to be included as separate variables in genetic risk modeling due to their independent risk contribution.^25^ On the other hand, including uninformative, non-causal variants in prediction models may introduce “noise” that obscures the effects of important variants. In a study by Dickson et al.,^26^ a model incorporating allelic *APOE* terms and just 20 additional Single-Nucleotide Polymorphisms (SNPs) outperformed the model that included thousands of SNPs in AD risk prediction (area under the receiver operating characteristic (AUROC): 0.75 vs. 0.63). Moreover, most current studies used longitudinal cohorts, which perform extensive testing and consensus criteria^27^ applied by clinicians with expertise in dementias to determine dementia diagnosis. While this approach ensures precision within research cohorts, it does not necessarily mirror the practicalities of real-world community settings. In real-world clinical care, the expertise in dementia may vary, and the criteria used for diagnosis may not always align with the stringent standards of research cohorts. Diagnoses documented in the Electronic Health Records (EHRs) capture these real-world data and, by routinely capturing patient data over extended periods, form an expansive longitudinal cohort ideal for real-world research. Compared to traditional cohorts, EHR cohorts offer additional benefits, such as vast sample sizes, diverse phenotypes, and a more inclusive representation of often underrepresented groups, like minorities and older adults.^28^ However, only a few genetic studies on dementia have been conducted within the context of EHR, and have predominantly focus on AD^11,29^

Finally, prior studies have primarily focused on the genetic risk prediction of AD. However, while AD accounts for a significant portion of dementia cases, concentrating solely on it risks overlooking the broader scope of cognitive disorders. In real-world scenarios, many dementia cases display mixed pathologies,^30,31^ with mixed dementia being a common occurrence ^32^. Addressing dementia as a whole, rather than exclusively focusing on AD, could better reflect the clinical landscape and lead to interventions and therapies that benefit a larger cohort of affected individuals.^33^

Unfortunately, dementia remains significantly underdiagnosed in real-world community settings. Research comparing diagnoses from real-world sources like Medicare claims or EHR to the gold standard diagnoses from longitudinal cohort studies reveals a sensitivity range of just 50-65%.^34–39^ Early detection of all-cause dementia with genetic modeling can empower healthcare providers to pinpoint the appropriate diagnostic processes, streamline care coordination, manage symptoms effectively, and begin suitable treatments. The above-mentioned limitations underscore the need for more refined methodologies to develop genetic risk models across diverse populations accurately.

In the present study, we hypothesized that the risk SNPs associated with dementia, and their corresponding weights, may vary across diverse populations, namely Amerindian, African, and East Asian genetic ancestry. We further proposed that the prediction performance of dementia phenotypes in non-European populations could be enhanced by identifying biological-meaningful SNPs followed by sparse machine learning models within each genetic ancestry group. Thus, we present a novel approach for assessing individual dementia genetic risks across diverse populations.

Our approach addresses the previously noted limitations through several innovative measures. Firstly, we utilized functional and biological information to prioritize SNPs based on GWAS results, thereby targeting causal SNPs with the highest likelihood of contributing to dementia risk. Secondly, we employed machine learning algorithms to select important genetic variants. Our method allows for the fine-tuning of models across different ancestry groups, offering a significant advantage for non-European populations that are often underrepresented in GWAS studies. Finally, we developed and validated our models within real-world EHR settings, focusing on predicting dementia as an encompassing condition. This innovative approach holds promise for enhancing our understanding of individual dementia genetic risks and promoting health equity in genetic research.

## 2 Methods

### 2.1 Data source

#### 2.1.1 UCLA ATLAS Community Health Initiative

Our discovery cohort for model development was derived from the biobank-linked EHR of the UCLA Health System.^40^ The UCLA ATLAS Community Health Initiative collects biosamples from participants of a diverse population. Upon obtaining patient consent, these biological samples undergo genotyping using a customized Illumina Global Screening Array.^41^ Detailed information regarding the biobanking and consenting procedures can be referenced in our previous publications.^42,43^ After the genotype quality control described below, there were 54,935 individuals with genotype and UCLA EHR data. As all genetic data and EHRs utilized in this study were de-identified, the study was deemed exempt from human subject research regulations (UCLA IRB# 21-000435).

#### 2.1.2 All of Us Research Hub

We validated our models and findings using All of Us Research Hub data. As one of the most diverse biomedical data resources in the United States, the All of Us Research Program serves as a centralized data repository, offering secure access to de-identified data from program participants.^44^ For our validation, we utilized data release version 7, encompassing 409,420 individuals, of which 245,400 have undergone whole genome sequencing.

### 2.2 Patient genetic data preprocessing

#### 2.2.1 Quality control

The quality control process was conducted using PLINK v1.9,^45^ adhering to established guidelines.^40^ We removed samples with a missingness rate exceeding 5%. Low-quality SNPs with >5% missingness and monomorphic and strand-ambiguous SNPs were excluded. Post-quality control, we performed genotype imputation via the Michigan Imputation Server.^46^ This step was crucial to augment the coverage of genetic variants and enable the comparison of results across diverse genotyping platforms. SNPs with imputation r^2^ <0.90 or MAF <1% were pruned from the data. After quality control measures and imputation, there were 21,220,668 genotyped SNPs across a sample of 54,935 individuals. Finally, we restricted our analyses to SNPs that overlapped between UCLA ATLAS and All of Us, amounting to a total of 8,705,988 SNPs. This approach ensured consistency in the genetic variables under consideration across both datasets.

#### 2.2.2 Inferring genetic ancestry

Genetic ancestry refers to the geographic origins of an individual’s genome, tracing back to their most recent biological ancestors while largely excluding cultural aspects of their identity.^47^ Genetic Inferred Ancestry (GIA) employs genetic data, a reference population, and inferential methodologies to categorize individuals within a group likely to share common geographical ancestors.^48^ In our UCLA ATLAS sample, we used the reference panel from the 1000 Genomes Project^49^ and principal component analysis^50^ to infer a patient’s genetic ancestry. GIA groups included European American (EA), African American (AA), Hispanic Latino American (HLA), East Asian American (EAA), and South Asian American (SAA). For instance, we designated individuals within the United States whose recent biological ancestors were inferred to be of Amerindian ancestry as “HLA GIA“.^51^ In addition, we calculated ancestry-specific principal components within each GIA group using principal component analysis.

### 2.3 Genetic predictors

#### 2.3.1 GWAS selection

Our study’s initial step is identifying potential risk SNPs as candidate predictors for dementia GWASs. A summary of the GWASs used and steps to select candidate SNPs in our study can be found in **Supplementary Table 1** and **Supplementary Figure 1**.

We selected GWASs for AD,^5,52,53^ PDD,^54^ PSP,^55^ LBD,^56^ and stroke^57^ phenotypes. For AD GWASs, we included three different GWASs conducted on diverse populations, including European,^5^ African American,^52^ and multi-ancestries.^53^ The summary statistics from all these GWAS are publicly available. Detailed information regarding the recruitment procedures and diagnostic criteria can be found in the original publications.

#### 2.3.2 Candidate SNPs identification and annotation

A significant proportion of GWAS hits are found in non-coding or intergenic regions,^58^ and given the correlated nature of genetic variants in Linkage disequilibrium (LD), distinguishing causal from non-causal variants often proves challenging based solely on association P-values from GWASs.^59^ Pinpointing the most likely relevant causal variants typically involves understanding the regional LD patterns and assessing the functional consequences of correlated SNPs, such as protein coding, regulatory, and structural sequences.^60^ Several functionally validated variants have been proved to be clinically relevant to the pathogenesis of diseases, as confirmed through in vitro or in vivo experimental validation.^61^ To address this, we utilized the Functional Mapping and Annotation of Genome-Wide Association Studies (FUMA), a tool that leverages information from biological data repositories and other resources to annotate and prioritize SNPs.^59^

For each GWAS summary statistic, we first identified genomic risk loci using a P-value threshold (<5e-8) and a pre-calculated LD structure (r^2^<0.2) based on the relevant reference population from the 1000 Genomes.^49^ Subsequently, we identified two distinct sets of SNPs:

1. **Independent genome-wide-significant SNPs**: We selected the SNP with the most significant GWAS P-value within each genomic risk locus. This process was iterated until all SNPs were assigned to a risk locus cluster or considered independent.
2. **Independent gene-annotated SNPs**: We prioritized SNPs based on their functional consequences on genes. In FUMA, the mapping from SNPs to genes was achieved by performing ANNOVAR^62^ using Ensembl genes (build 85). SNPs were mapped to genes through positional mapping, eQTL associations, and 3D chromatin interactions. The Combined Annotation-Dependent Depletion (CADD) score^63^ was used to select potential causal SNPs, with the SNP possessing the highest CADD score within each genomic risk locus being chosen, indicating a higher probability of the variant being deleterious.

The identified independent genome-wide-significant SNPs and independent gene-annotated SNPs were subsequently used in constructing the disease PRSs and as candidate features in dementia prediction models. To ensure the robustness of our findings, we also adopted a stringent r^2^ cut-off (<0.1) to define independent genome-wide-significant SNPs, ensuring the selected SNPs were independent.

#### 2.3.3 Polygenic risk scores and *APOE-ε4*

We computed the disease-specific PRS as the sum of an individual’s risk allele dosages, each weighted by its corresponding risk allele effect size from the GWAS summary statistics, as shown in the PRS equation 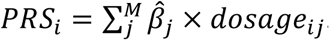. All PRSs were then standardized to a mean of 0 and a standard deviation of 1. The standardization process used the 1000 Genome European genetic ancestry as the reference population, ensuring that the scores’ range and values are comparable across different GWASs. For each phenotype, we employed two distinct sets of SNPs identified by FUMA, namely the independent genome-wide-significant SNPs and independent gene-annotated SNPs, to calculate two respective PRSs: *PRS.psig* and *PRS.map*. The *APOE* gene has two variants, rs7412 and rs429358, which determine the three common isoforms of the apoE protein: E2, E3, and E4, encoded by the ε2, ε3, and ε4 alleles.^64^ Previous research has demonstrated that out of the three polymorphic forms of *APOE*, carriers of *APOE-e4* are at a higher risk of developing AD, and this association exhibits a dose-dependent effect.^65^ Therefore, to quantify the *APOE* genotype in our study, we created a numerical variable, “*APOE*-*e4count*“, with the two variants mentioned above, representing the number of ε4 alleles (0, 1, or 2) carried by each individual.

### 2.4 Dementia definition and demographic features

The primary outcome of interest was dementia, which we defined using the ICD-10 codes (**Supplementary Table 2)**. The demographic variables considered in our study were self-reported sex and age. The age of each participant, measured in years, was calculated based on their self-reported birth date and the dates of their encounters. For individuals diagnosed with dementia, we determined the age at dementia onset.

### 2.5 Analytical sample selection

To focus on patients with longitudinal records, our analyses included patients with complete demographic data (age and sex) who had at least two medical encounters after age 55. We also applied a restriction of age at the last recorded encounter to be less than 90 as patients in the UCLA EHR dataset are censored when older than 90.

We identified eligible dementia cases as patients with at least one encounter with a recorded dementia diagnosis, provided that the initial onset of the condition occurred after age 55. To qualify as an eligible control, subjects were required to meet the following criteria: 1) not have any recorded dementia or related diagnoses, as determined by a set of predefined exclusion phenotypes;^66^ 2) age at the last recorded visit >=70, to exclude younger patients who may not have manifested signs of dementia; and 3) a minimum of five years’ length of records with an average of at least one encounter per year, thereby minimizing the potential for bias associated with misdiagnosis.

Upon the application of these selection criteria, the resultant sample served as the pool for permutation resampling and subsequent modeling in our study.

### 2.6 Prediction of dementia risk with machine learning models

In our discovery study, we developed a series of logistic regression models to predict the binary dementia phenotype in the UCLA ATLAS sample, stratified by GIA groups.

#### 2.6.1 Permutation resampling

In order to fortify the reliability of our findings, we employed the permutation resampling methodology to assess model performance, ascertain feature importance, and evaluate statistical significance. Specifically, we conducted random sampling from the pool of eligible controls, maintaining a case-to-control ratio of 1:3, and utilized the amalgamated case and control samples for the following modeling process. This iterative procedure was repeated 1000 times.

#### 2.6.2 Regress out demographic variable effects

To distinctly assess genetic influences, our analysis commenced by mitigating the impact of demographic factors, encompassing age, sex, and ancestry-specific principal components (PCs), from the predictive model. We first employed a logistic regression model that exclusively utilized these variables to predict dementia status. Subsequently, we derived the predicted values for each patient through this model. Applying an appropriate inverse link function (e.g., logit), we then subtracted these predicted values from the ultimate outcome (dementia status), generating an “offset” value. These offset values encapsulated the dementia status, after regressing out the effects of demographic variables and genetic population structure.

#### 2.6.3 Genetic prediction models

Next, we trained genetic risk models to predict the outcome (dementia status) with the offset corrections applied in the linearized space, i.e., *y^*_*i*_ = *g*^−1^(*β*_0_ + *β*_1_*x*_*i*1_ + ⋯ + *β*_*p*_*x*_*ip*_ + *offset*_*i*_ ), where *y^*_*i*_ represents the predicted dementia status, and *g*^−1^(*·*) is the inverse of the link function.^67^ We compared four different sets of predictors: 1) *APOE* status, 2) AD PRS, 3) multiple PRSs, and 4) smaller SNP sets with Elastic Net regularization. The latter involved the application of a regularization technique known as Elastic Net to smaller sets of SNPs.^68^ For multiple PRS models, we crafted models utilizing diverse AD PRSs of varying ancestries or PRSs derived from other GWASs focused on neurodegenerative diseases. Across all models, we employed a 5-fold cross-validation methodology to authenticate their predictive efficacy, with the final results reported on the combined hold-out testing set.

The primary assessment criterion was the Area Under the Precision-Recall Curve (AUPRC), specifically chosen for its appropriateness in scenarios involving imbalanced datasets where the number of cases is significantly outnumbered by controls.^69^ Additionally, the AUROC was reported as a comprehensive metric for model evaluation. To determine the optimal threshold, we selected the point that maximized the Matthews Correlation Coefficient (MCC).^28^ Subsequent performance metrics, such as the F1 score, accuracy, precision, recall, and specificity, were computed based on this threshold. The 95% confidence intervals (CIs) and p-values 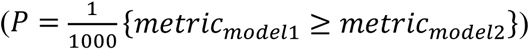 were derived through 1000 permutations as described previously.

### 2.7 Validations in the All of Us sample

We conducted a validation study using the All of Us cohort to assess the generalizability of our findings derived from the UCLA ATLAS sample. We selected a comparable sample from the All of Us Research Hub, adhering to the same criteria and sampling scheme for the GIA groups in the UCLA ATLAS sample. The same methodologies were employed to define dementia cases and controls. We extracted the same genetic risk loci from the All of Us Whole Genome Sequencing data for PRS construction or those identified through Elastic Net models in the UCLA ATLAS sample. We employed a consistent methodology to regress out demographic variables and genetic population structure (i.e., PCs) as a preliminary step. This approach was undertaken to derive offset corrections, mirroring the procedures employed in our prior research. By regressing out these factors, we aimed to ensure that the statistical models accurately reflect the intrinsic genetic associations, unconfounded by extraneous demographic or population structure influences.

We compared three models in the All of Us sample: 1) the *APOE-e4* model; 2) the best-performing PRS model; and 3) the best-performing Elastic Net SNP model. The same evaluation metrics were utilized for model comparisons.

### 2.8 Gene mapping and gene set analysis

To facilitate biological interpretations, we employed FUMA’s positional, eQTL, and chromatin interaction mapping to associate dementia risk SNPs, identified from the top-performing Elastic Net SNP models, with specific genes.^59^ We then tested these mapped genes against gene sets procured from MsigDB, such as positional gene sets and Gene Ontology (GO) gene sets, to assess the enrichment of biological functions through hypergeometric tests. To correct for multiple testing, we implemented the Benjamin-Hochberg adjustment.^70^ Using heatmaps, we reported and visualized gene sets with an adjusted P-value ≤0.05 and more than one overlapping gene.

## 3 Results

### 3.1 Sample description

The study’s primary dataset for model development was derived from EHR linked to the biobank of the UCLA Health System.^40^ A detailed depiction of the sample selection steps and resampling scheme is provided in **Figure 1A**.

**Figure 1.**
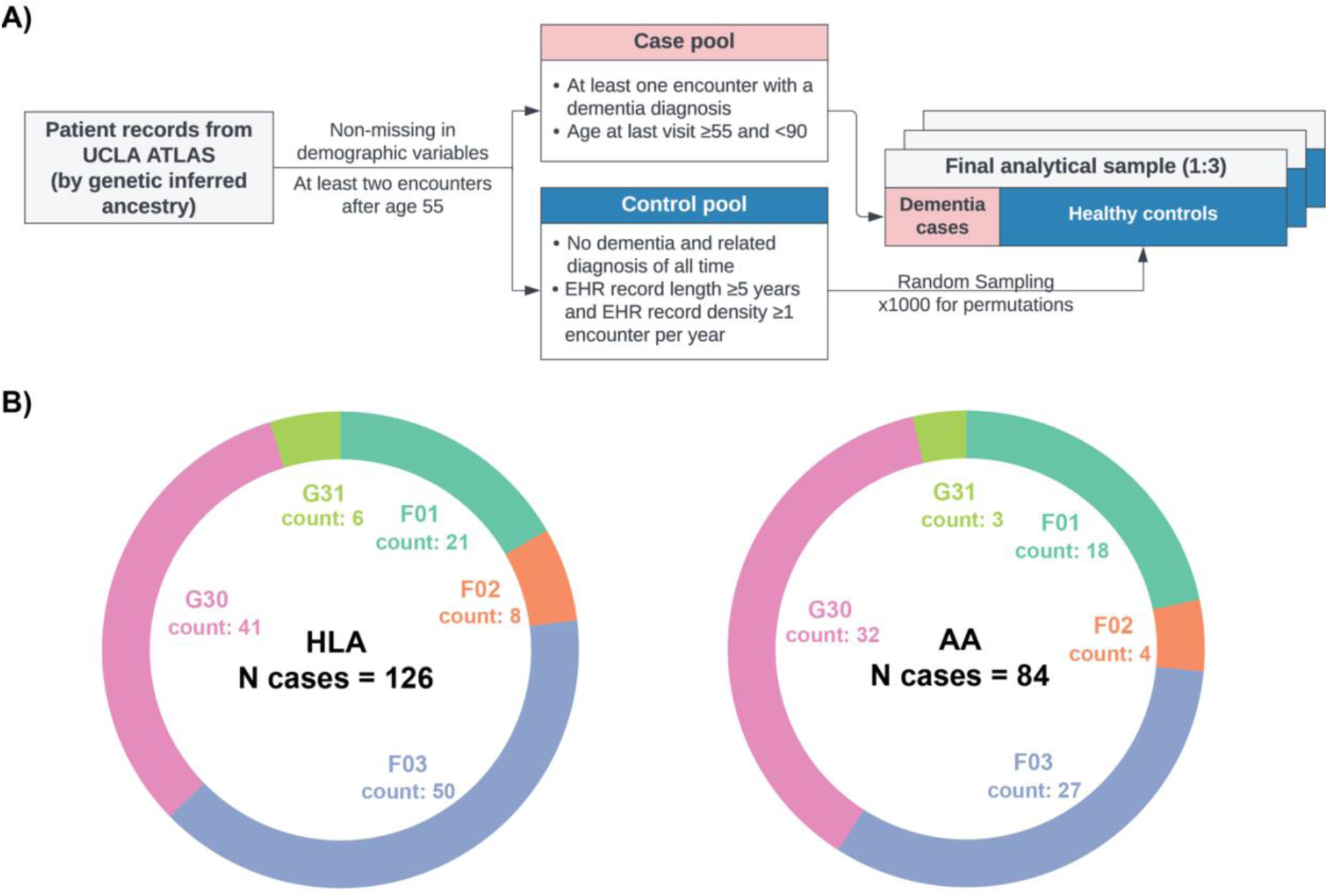
Sample selection steps and dementia patient characteristics by genetic inferred ancestry groups, UCLA ATLAS sample. A) Inclusion criteria and case-control selection steps. B) Distribution of diagnosis in ICD-10 codes by genetic inferred ancestry groups. *Abbreviations: AA, African Americans; HLA: Hispanic Latino Americans. ICD-10 codes descriptions: G30, Alzheimer’s disease; F03, Unspecified dementia; F02, Dementia in other diseases classified elsewhere; F01, Vascular dementia; G31, Other degenerative diseases of nervous system, not elsewhere classified*.

Figure 1B illustrates the finalized UCLA ATLAS samples, stratified by GIA groups. Notably, the HLA sample comprised 610 patients, while the AA sample consisted of 440 patients, with 126 and 84 dementia cases, respectively, within each group. The distribution of International Classification of Diseases, 10th Revision (ICD-10) diagnosis codes remained relatively consistent across the two GIA samples, with Alzheimer’s disease (G30) and unspecified dementia (F03) being the most prevalent diagnoses. However, it is important to highlight that the AA group exhibited a higher proportion of patients diagnosed with vascular dementia (F01) compared to the HLA group. The EAA group, with a limited case count (N = 75), was excluded from primary analyses but included in sensitivity analyses.

Within each GIA group, we found that eligible controls, due to the more stringent inclusion criteria, displayed a longer span of records and more encounters. There were no significant differences in other EHR features between dementia cases and controls (**Table 1**).

**Table 1.** Descriptive statistics of demographic and electronic health record features by case/control groups, UCLA ATLAS sample, stratified by genetic inferred ancestry group.

|  | Hispanic Latino Americans (N = 610) |  |  | African Americans (N = 440) |  |  |
| --- | --- | --- | --- | --- | --- | --- |
|  | Cases | Controls | P value | Cases | Controls | P value |
| N | 126 | 484 | - | 84 | 356 | - |
| Age | 78.4 (71.3, 81.7) | 75.3 (72.6, 79.6) | 0.2 | 78.0 (70.1, 82.6) | 75.7 (72.7, 79.9) | 0.7 |
| Sex (Female) | 72 (57%) | 300 (62%) | 0.30 | 46 (55%) | 218 (61%) | 0.30 |
| Span of records (in yrs) | 5.9 (2.8, 8.8) | 9.6 (7.7, 10.9) | <0.001* | 6.2 (3.1, 10.1) | 9.9 (8.1, 11.4) | <0.001* |
| Encounters per year | 16 (7, 25) | 14 (8, 20) | 0.05 | 14 (6, 28) | 13 (9, 21) | 0.60 |
| Number of encounters | 73 (26, 156) | 124 (73, 205) | <0.001* | 65 (28, 183) | 140 (84, 210) | <0.001* |
| Number of unique diagnosis | 68 (36, 113) | 71 (47, 108) | 0.40 | 61 (41, 99) | 73 (47, 103) | 0.20 |

**Notes:** Continuous variables were reported as median (IQR), and categorical variables were reported as n (%). P-values were calculated based on Wilcoxon rank sum test or Pearson's Chi-squared test as appropriate. \* Statistically significant at level 0.05.

### 3.2 Performance comparison for dementia phenotype prediction task

We developed and evaluated a series of logistic regression models to predict the binary dementia phenotype within the UCLA ATLAS sample, stratified by GIA groups. After regressing out the effects of age, sex, and ancestry-specific genetic variations as represented by PCs, we constructed genetic risk models for dementia, incorporating offset corrections within a linearized framework. The predictive capabilities of these models were assessed using four distinct sets of genetic markers: 1) *APOE-e4* counts, 2) AD PRS, 3) a composite of multiple PRSs, and 4) select SNPs refined through Elastic Net regularization.^68^ For the selection of SNP sets, we utilized the FUMA tool^59^ to prioritize independent genome-wide-significant SNPs or independent gene-annotated SNPs. We employed the permutation resampling methodology (1000 times) to assess model performance, ascertain feature importance, and evaluate statistical significance (details see **Methods**).

The overall performances of models for predicting dementia phenotypes are visually represented in Figure 2. No discernible differences were observed among *APOE-e4* and all PRS models, irrespective of the SNP set employed for PRS construction—whether derived from ancestry-specific GWASs, genome-wide-significant SNPs, or gene-annotated SNPs. Notably, the predictive performance of *APOE-e4* and all PRS models within the AA GIA sample exhibited inferior outcomes compared to the HLA GIA sample, particularly evident in the AUPRC.

**Figure 2.**
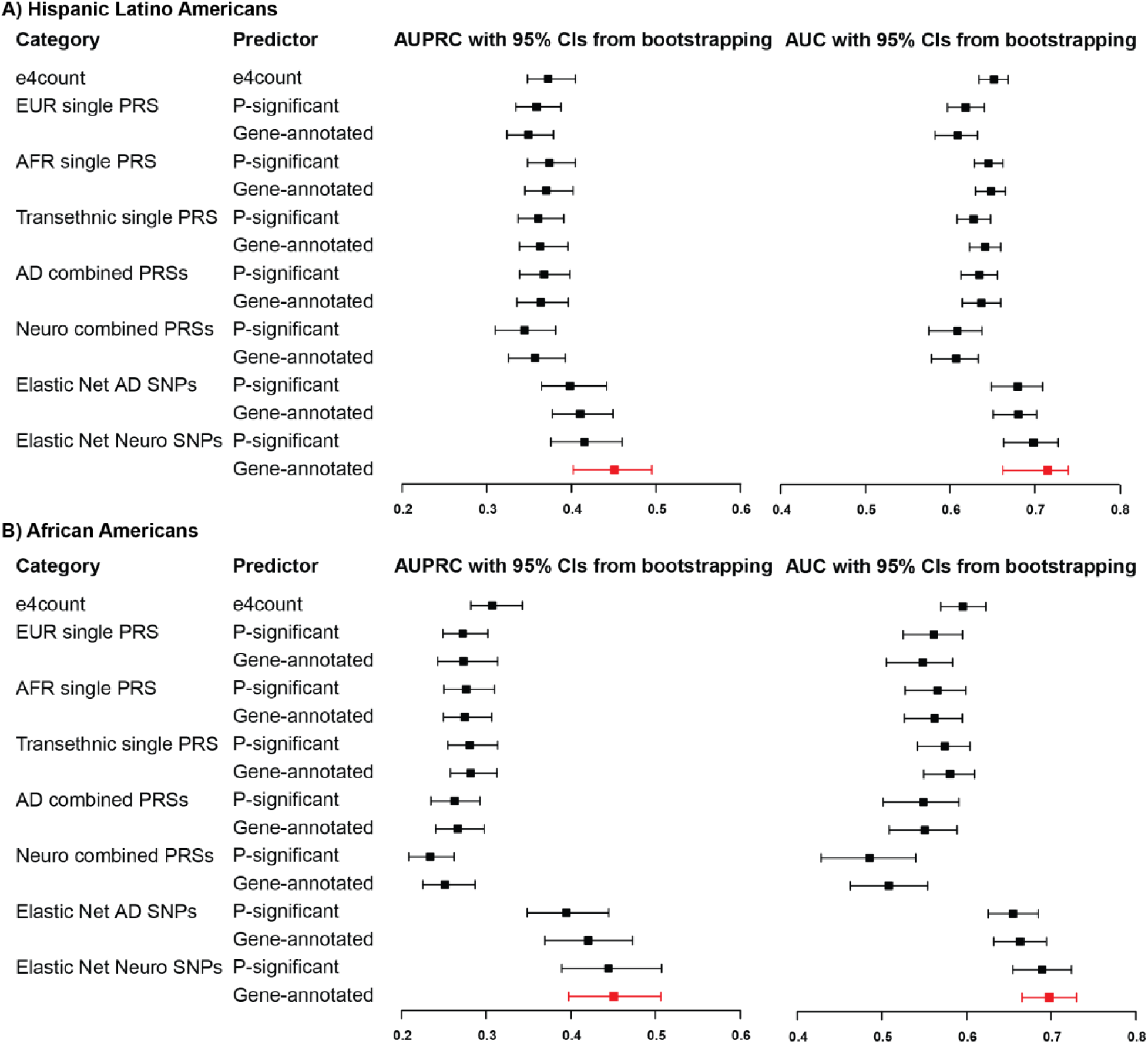
Overall model performance of *APOE-e4* count, polygenic risk score, and Elastic Net SNP models in dementia genetic prediction, UCLA ATLAS sample, stratified by genetic inferred ancestry group. All models (if not other specified) have regressed out age, sex, and ancestry-specific principal components. *Abbreviations: AD, Alzheimer’s Disease; AUROC, Area Under the ROC Curve; AUPRC, Area Under the Precision-Recall Curve; EUR, European; PRS, Polygenic Risk Score; SNP, Single-Nucleotide Polymorphism*.

Elastic Net SNP models demonstrated an overall improvement in dementia prediction across both GIA groups. The model incorporating gene-annotated SNPs from AD and other dementia-related disease GWASs emerged as the most effective, indicating a collective contribution from SNPs associated with various dementia-related diseases. Specifically, the leading Elastic Net SNP model for HLA GIA sample significantly enhanced the AUPRC by 22% (0.451 vs. 0.371, p-value = 0.003), and the AUROC by 11% (0.715 vs. 0.648, p-value = 0.008) compared to the best PRS model. Furthermore, this model outperformed the *APOE-e4* count model, with increments of 21% in AUPRC (p-value = 0.003) and 10% in AUROC (p-value = 0.007). This model’s efficacy was even more pronounced within the AA GIA sample, with an increase in AUPRC by 61% (p-value < 0.001) and the AUROC by 21% (p-value < 0.001) in comparison to the best PRS model. Relative to the *APOE-e4* count model, the improvements were 47% in AUPRC (p-value < 0.001) and 17% in AUROC (p-value < 0.001).

We also noted a substantial enhancement in the other performance metrics (based on the threshold that maximized the MCC) of the Elastic Net SNPs models compared to other models across both GIA samples (**Supplementary Table 3**). This was evidenced by marked improvements in accuracy, precision, and the F1 score. In our sensitivity analysis, applying a more stringent r^2^ cut-off (<0.1) for defining independent genome-wide-significant SNPs yielded results consistent with our initial findings, as detailed in **Supplementary Table 4**. In summary, models leveraging SNPs as features identified through machine learning methods possess the potential to surpass those relying solely on summary scores such as PRSs. Furthermore, selecting SNPs mapped to genes using functional genomic data holds promise for further refining predictive performance.

### 3.3 Featured risk variants and mapped genes

In our analysis of the best-performing Elastic Net SNPs models, we further examined the features selected by each model. The HLA and AA models identified 15 and 10 risk SNPs, respectively. A detailed list of SNPs, including related information, is provided in **Table 2**.

**Table 2.**
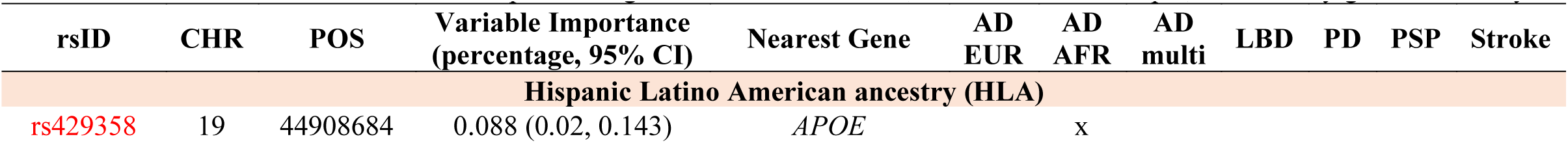
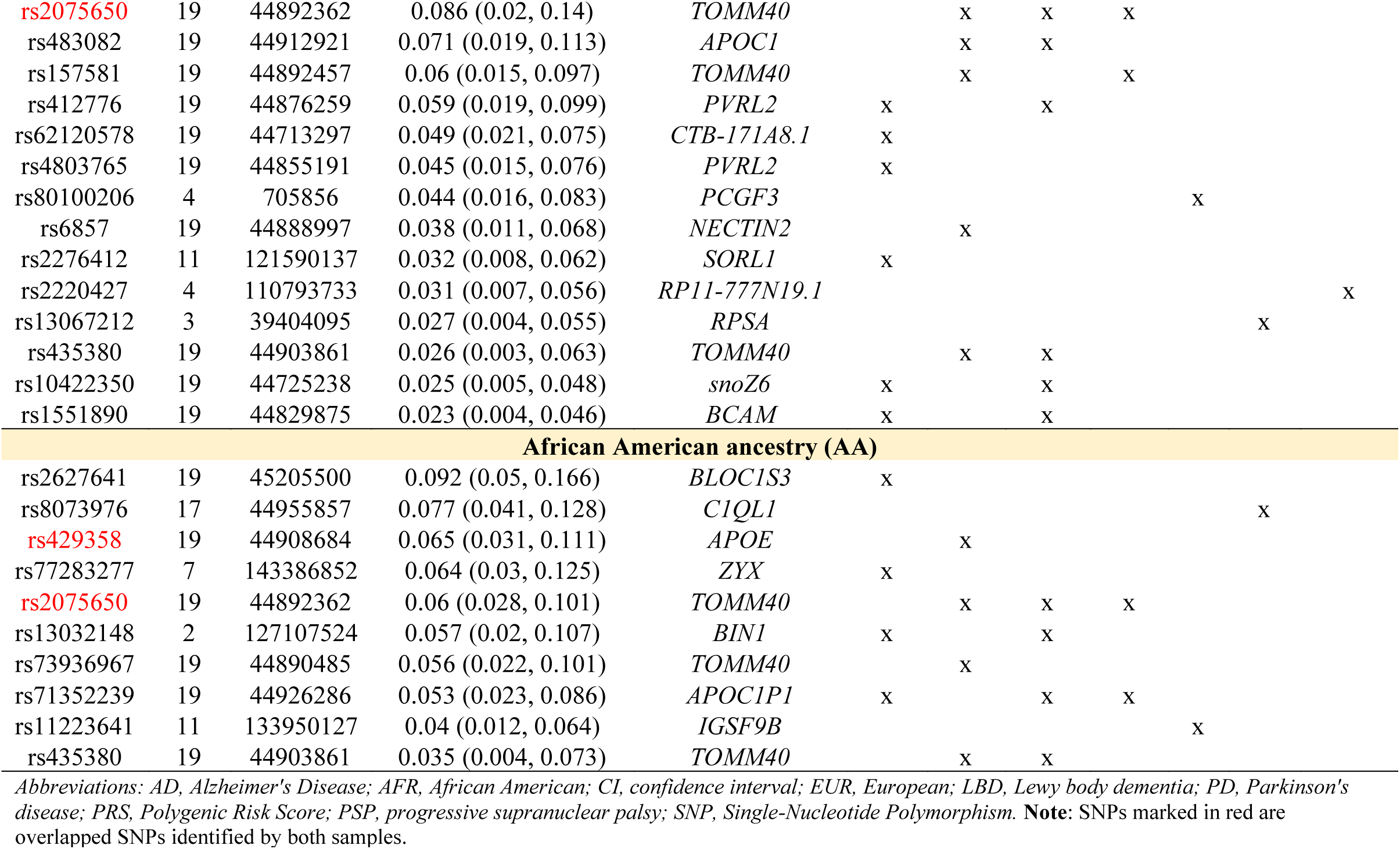
Featured risk SNPs from the best-performing Elastic Net SNP model, UCLA ATLAS sample, stratified by genetic ancestry.

By assessing the feature importance of the SNPs chosen by the models, we discovered that rs429358 (chr19:44908684, nearest gene: *APOE*), rs2075650 (chr19:44892362, nearest gene: *TOMM40*), and rs483082 (chr19: 44912921, nearest gene: *APOC1*) were selected as the top three important predictor for the HLA GIA group, together accounting for ∼25% of the total predictive importance. Conversely, for the AA GIA group, the most influential predictors were identified as rs2627641 (chr19:45205500, nearest gene: *BLOC1S3*), rs8073976 (chr17:44955857, nearest gene: *C1QL1*), and rs429358 (chr19:44908684, nearest gene: *APOE*). Two AD-associated risk SNPs, rs429358 and rs2075650, were pinpointed by both GIA Elastic Net SNPs models, albeit with slight variations in their relative importance. Moreover, both models identified several risk SNPs of PDD and progressive supranuclear palsy (PSP) as crucial predictors of dementia. However, there were notable differences between the models. For instance, the AA GIA model ascribed significant importance to a PSP-associated risk SNP, rs8073976, located on chromosome 17. Interestingly, stroke-risk SNPs were only identified as important predictors by the HLA GIA model, underscoring the distinct genetic underpinnings influencing these different ancestry groups.

To better understand the biological functions and pathways associated with the identified risk variants, we then mapped those featured risk SNPs to genes. This was also achieved using FUMA, which incorporates positional, eQTL, and 3D chromatin mapping.^59^

Notably, four genes were identified by both non-European GIA models (Figure 3 **& Supplementary Table 5**). All shared genes were located near *chr19q13*, which includes the well-established AD risk gene cluster, *APOE-TOMM40-APOC1.*^71^ According to the enrichment analysis results, these shared genes are predominantly involved in biological pathways associated with lipid metabolism. These pathways encompass processes such as the assembly and organization of protein-lipid complexes, as delineated by the GO terms. Additionally, these genes play an essential role in regulating cholesterol, triglyceride, amyloid proteins, and lipoprotein particles, further underscoring the significance of lipid metabolic processes in dementia. In addition, we investigated ancestry-specific genes. For instance, genes near the *chr17q21* (e.g., *CCDC43, GFAP, and C1QL1*), and the *chr11q25* region (e.g., *GSF9B* and *JAM3*) were uniquely pinpointed by the AA GIA model.

**Figure 3.**
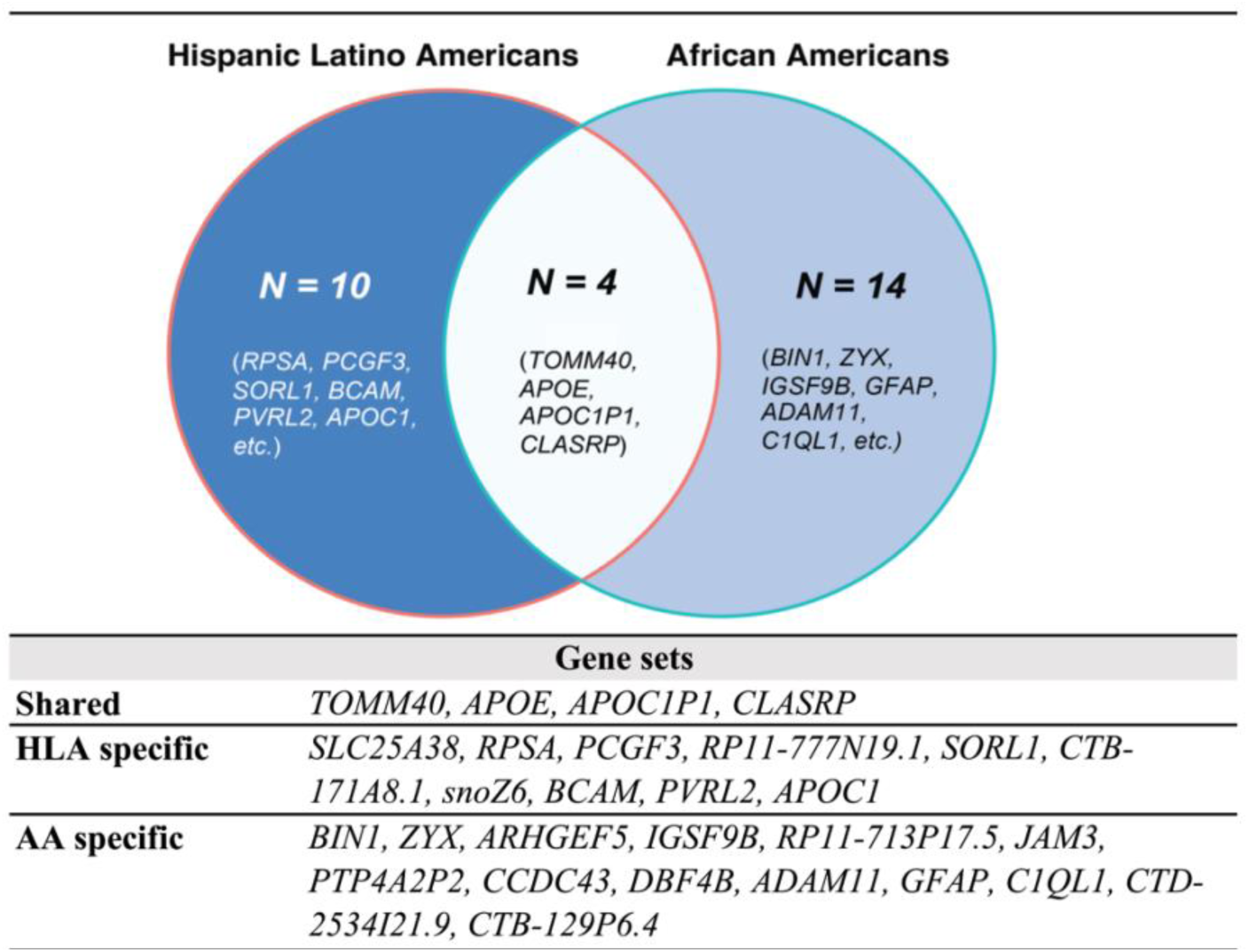
Shared and ancestry-specific risk genes identified by the best-performing Elastic Net SNP models, UCLA ATLAS sample.

In the sensitivity analyses, we performed dementia risk modeling in the EAA GIA sample (N = 673). Similar to other GIA groups, the model incorporating gene-annotated SNPs from AD and other dementia-related disease GWASs performed the best compared to all other models, enhancing the AUPRC by 11% (0.511 vs. 0.459), and the AUC by 7% (0.754 vs. 0.703) compared to the best PRS model. Despite these improvements, the differences in performance between the leading Elastic Net SNP model and other models did not reach statistical significance (AUPRC: p-value = 0.438; AUROC: p-value = 0.376). Among the featured 12 risk SNPs, rs429358 (chr19:44908684, nearest gene: *APOE*), rs35106910 (chr19:44781009, nearest gene: *CBLC*), and rs66626994 (chr19:44924977, nearest gene: *APOC1P1*) were the most significant predictors for the EAA GIA group, collectively accounting for ∼32% of the overall predictive importance. After mapping featured SNPs to gene, we also identified the AD-risk gene cluster, *APOE-TOMM40-APOC1,* as well as the gene region near *chr17q21* (e.g., *FMNL1 and SPPL2C*) (**Supplementary Table 6A-D**).

### 3.4 Validations in the All of Us sample

We conducted a validation study using the All of Us cohort to evaluate the broad applicability of our findings obtained from the UCLA ATLAS sample. A comparable sample was selected from the All of Us Research Hub, employing the same selection scheme to their corresponding GIA groups in the UCLA ATLAS sample. However, due to the limited number of eligible dementia cases (N case = 8) in the All of Us EAA GIA sample, we could only validate our models and findings in the HLA (N_case = 81, N_control = 445) and AA (N_case = 181, N_control = 2,463) samples. In contrast to the UCLA ATLAS samples, the All of Us cohort samples exhibited a younger demographic profile, with participants having comparatively shorter durations of EHR documentation and fewer recorded healthcare visits. Within each GIA sample, we found similar distributions of demographics and EHR features between dementia cases and eligible controls (**Supplementary Table 7-8**).

We applied the model weights trained from the UCLA ATLAS sample to the All of Us sample, stratified by GIA groups. In the comparison of three representative models, namely 1) the *APOE-e4* model; 2) the best-performing PRS model; and 3) the best-performing Elastic Net SNP model, our results mirrored those from the UCLA ATLAS sample, with the Elastic Net SNP model, which included gene-annotated SNPs from GWASs of AD and other dementia-related diseases, outperforming all other models in terms of the AUPRC and AUC in both the HLA and AA GIA samples (**Table 3**).

**Table 3.**
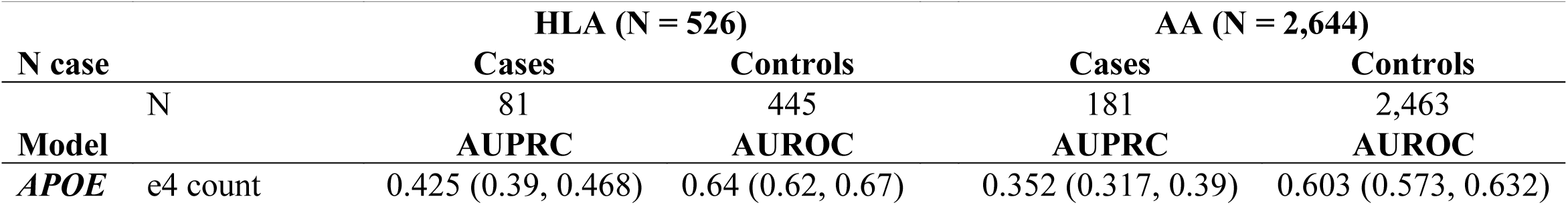
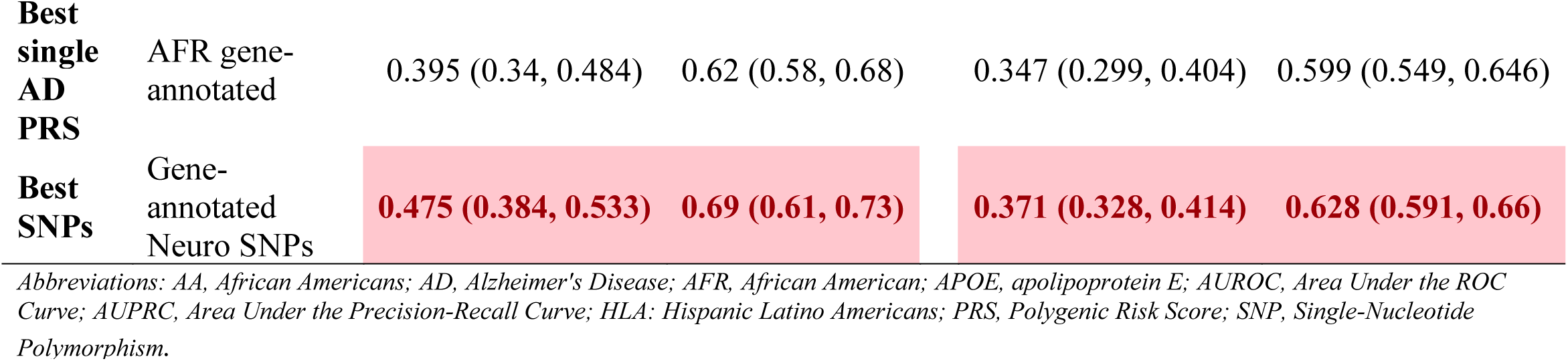
Overall model performance of *APOE-e4* count, polygenic risk score, and Elastic Net SNP models in dementia genetic prediction in validation of All of Us sample, stratified by genetic inferred ancestry.

|  |  | HLA (N = 526) |  | AA (N = 2,644) |  |
| --- | --- | --- | --- | --- | --- |
| N case |  | Cases | Controls | Cases | Controls |
|  | N | 81 | 445 | 181 | 2,463 |
| Model |  | AUPRC | AUROC | AUPRC | AUROC |
| *APOE* | e4 count | 0.425 (0.39, 0.468) | 0.64 (0.62, 0.67) | 0.352 (0.317, 0.39) | 0.603 (0.573, 0.632) |
| **Best single AD PRS** | AFR gene-annotated | 0.395 (0.34, 0.484) | 0.62 (0.58, 0.68) | 0.347 (0.299, 0.404) | 0.599 (0.549, 0.646) |
| **Best SNPs** | Gene-annotated<br>Neuro SNPs | **0.475 (0.384, 0.533)** | **0.69 (0.61, 0.73)** | **0.371 (0.328, 0.414)** | **0.628 (0.591, 0.66)** |

Abbreviations: AA, African Americans; AD, Alzheimer's Disease; AFR, African American; APOE, apolipoprotein E; AUROC, Area Under the ROC Curve; AUPRC, Area Under the Precision-Recall Curve; HLA: Hispanic Latino Americans; PRS, Polygenic Risk Score; SNP, Single-Nucleotide Polymorphism.

In particular, the Elastic Net SNP model demonstrated a substantial improvement in the AUPRC, outperforming the *APOE-e4* model by 12% in AUPRC (p-value = 0.082), and the best AD PRS model (AD AFR *PRS.map*) by 20% in AUPRC (p-value = 0.034) in the HLA GIA sample. Similarly, in the AA GIA sample, the Elastic Net SNP model showed an enhancement of 5.4% (p-value = 0.083) and 6.9% (p-value = 0.528) in the AUPRC over the *APOE-e4* and best AD PRS model, respectively.

## 4 Discussion

Traditional genetic risk models have faced limitations in effectively capturing causal disease risk variants and accurately assessing genetic risks across diverse populations. To address these challenges, our present study introduces a novel approach to predicting dementia risks by leveraging functional mapping of genetic data in conjunction with machine learning methods in the real-world EHR setting. Our proposed method shows remarkable improvements in prediction performance compared to well-known approaches like *APOE* gene and PRS models. We successfully identified shared and ancestry-specific risk genes and biological pathways contributing to dementia risks for each non-European GIA group. Finally, we bolstered the reliability and generalizability of our findings by validating our models using a comparable EHR sample from the All of Us cohort.

Our study highlights the significance of prioritizing biologically meaningful SNPs in genetic prediction. GWASs often identify genomic regions with multiple correlated SNPs, which may encompass several closely located genes. However, not all of these genes are relevant to the disease.^72^ Functional annotation of genetic variants enabled us to target potential causal SNPs by considering various factors, such as regional LD patterns, functional consequences of variants, their impact on gene expression, and their involvement in chromatin interaction sites.^59^ In our models developed on UCLA ATLAS samples, we achieved significant improvements in model performance by prioritizing biologically meaningful SNPs, ranging from 21-61% in AUPRC and 10-21% in AUROC across different GIA groups, compared to the *APOE-e4* count and the best-performing PRS models. These results underscore the critical role of considering functional and biological information in enhancing the performance of genetic prediction models, especially in diverse populations.

It is worth highlighting that no discernible performance differences were observed between PRSs constructed using genome-wide-significant and gene-annotated SNPs. This can be attributed to the strong LD between genome-wide-significant and gene-annotated SNPs within the same genomic region. As a result, these SNPs tend to have similar effect estimates in the GWASs.

Thus, it is expected that the PRSs built with these two sets of SNPs would exhibit a high correlation (**Supplementary Table 9**), which further supports the notion that the choice of genome-wide-significant or gene-annotated SNPs does not significantly impact the predictive performance of the PRSs in our study.

Moreover, our study emphasizes the significance of incorporating risk factors from multiple dementia-related diseases when developing predictive models for complex conditions like dementia. Both ancestry-specific Elastic Net SNP models highlighted several PD and PSP risk variants as significant predictors of dementia. This finding aligns with the well-known complexity of dementia as a multifactorial disorder that shares common features with these related conditions.^73^ However, it is worth noting that including PRSs of those diseases did not significantly improve the overall performance (Figure 2). This result is consistent with research conducted by Clark et al.,^74^ in which they demonstrated that a combined genetic score, which incorporated risk variants for AD and 24 other traits, had an equivalent predictive power as the AD PRS on its own. One possible explanation is that many traits were not dementia etiologies and diluted the effects of the true causal SNPs in the models.

Our proposed Elastic Net SNPs models identified several shared risk factors across different ancestries. Notably, a substantial proportion of the identified shared genes were found near the *chr19q13* region, which is well-known for the AD risk gene cluster comprising *APOE-TOMM40-APOC1*. These findings align with previous research,^6,52,64^ further supporting the significance of this genomic region in contributing to the genetic risks associated with dementia. At the same time, we have discovered compelling evidence supporting our hypothesis that risk SNPs associated with dementia, along with their corresponding weights, exhibit significant variations across diverse populations. Notably, our analysis of PRS models revealed that the performance of PRS built with the European population GWAS was worse when predicting a non-European GIA group. On the other hand, we also observed that the *APOE-e4* count model performed better than most PRS models in HLA and AA GIA samples. These finding further reinforces the limitations of standard PRS when applied to non-European populations, in which attempting to transfer GWAS effect size from one GIA to another GIA, or when using matched genetic ancestry GWAS with smaller sample size, as demonstrated in several AD and other phenotype studies.^75–78^

In addition, we observed notable differences in the feature importance of various SNPs within the best-performing Elastic Net models across distinct GIA groups. Consequently, this led us to identify ancestry-specific genes and distinct biological pathways implicated in the genetic predisposition to dementia in diverse ancestral samples. These findings highlight the uniqueness of genetic risk factors and functional pathways in diverse population groups.

Finally, we validated our models using samples from separate EHR linked with genetic data (All of Us). Our proposed Elastic Net SNP model consistently outperformed the *APOE-e4* and the best PRS models. While the Elastic Net SNP model demonstrated effective performance in both HLA and AA populations, we observed a decrease in the general performance and significance (AUPRC and AUROC) in the All of Us sample compared to the UCLA ATLAS sample, particularly in the AA samples. One potential explanation for this discrepancy is the distinct population structure within each sample, as revealed by comparing patient characteristics (**Supplementary Table 7**). These findings underscore the influence of population-specific factors on the generalizability of genetic risk models, highlighting the critical need to account for population diversity in predictive models for complex diseases.

Our study boasts several notable strengths that contribute to its significance and impact. Firstly, machine learning techniques applied in our study allowed us to infer crucial dementia risk factors for underrepresented populations, such as HLA and AA, with GWAS summary statistics from extensively studied populations like Europeans. This approach enabled a deeper understanding of the genetic landscape of dementia in underrepresented populations, particularly valuable given the current limitations in large-sample-size GWASs specific to these groups. Secondly, we fortified the robustness and generalizability of our findings through the validation of our model on an independent dataset from the All of Us cohort. Furthermore, our innovative approach, which incorporated biologically relevant genetic markers and functional annotations, significantly enhanced the accuracy of disease prediction. This approach can be readily adapted to predict other complex diseases, extending the scope of its applications and enriching our understanding of diverse human populations’ genetic traits.

However, we acknowledge certain limitations. Firstly, we observed variations in the composition of dementia subtypes among different GIA groups’ case samples. Consequently, the distinct genes and biological pathways identified by different ancestry models should be interpreted with this consideration. Secondly, although our study identified potential risk SNPs and genes associated with dementia, additional experimentation is necessary to understand the precise mechanisms underlying the association of these factors with dementia. Thirdly, due to the limited number of dementia cases in the All of Us EAA GIA sample after applying our inclusion criteria, we could only validate our models and findings in the HLA and AA samples. As a result, the generalizability of our findings to the EAA ancestry is constrained.

In light of these limitations, further research with more extensive and diverse datasets, encompassing a broader range of dementia subtypes and GIA groups is imperative to strengthen the validity and applicability of our study’s outcomes. Such efforts will contribute to a more comprehensive understanding of the genetic complexities underlying dementia across diverse populations.

## 5 Conclusions

Our study introduces a novel and robust approach to assessing individual genetic risks for dementia across diverse populations in a real-world setting. Our study demonstrates the importance of considering functional and biological information and population diversity when developing predictive models for complex diseases like dementia. The findings from our research provide valuable insights into the intricate genetic factors underlying dementia. Moreover, this work opens up promising avenues for developing more accurate and efficient predictive models for complex genetic traits in diverse human populations. Such advancements can potentially be paired with the development of targeted treatments tailored to the specific genetic profiles of individuals affected by dementia and related conditions.

## 6 List of abbreviations

AA: African American
AD: Alzheimer’s disease
APOE: Apolipoprotein E
AUPRC: Area Under the Precision-Recall Curve
AUROC: area under the receiver operating characteristic
CADD: Combined Annotation-Dependent Depletion
CI: confidence intervals
EA: European American
EAA: East Asian American
EHR: Electronic Health Records
FTD: Frontotemporal dementia
FUMA: Functional Mapping and Annotation of Genome-Wide Association Studies
GIA: Genetic Inferred Ancestry
GO: Gene Ontology
GWAS: Genome-Wide Association Studies
HLA: Hispanic Latino American
LBD: Lewy body dementia
LD: Linkage disequilibrium
MCC: Matthews Correlation Coefficient
PC: principal components
PDD: Parkinson’s disease dementia
PRS: Polygenic risk scores
SAA: South Asian American
SNP: Single-Nucleotide Polymorphisms

## 7 Declarations

### 7.1 Ethics approval and consent to participate

All human subjects involved in this study provided informed consent, ensuring their understanding and voluntary participation in the research.

### 7.2 Consent for publication

Not applicable.

### 7.3 Availability of data and materials

The Genome-Wide Association Study summary statistics data analyzed in this study are publicly available. Individual electronic health record data are not publicly available due to patient confidentiality and security concerns. Collaboration with the study authors who have been approved by UCLA Health for Institutional Review Board-qualified studies are possible and encouraged. Code is available on GitHub: https://github.com/TSChang-Lab/Dementia-prediction. Requests for additional information can be directed to the Lead Contact: Timothy S

Chang.

### 7.4 Competing interests

The authors declare that the research was conducted in the absence of any commercial or financial relationships that could be construed as a potential conflict of interest.

### 7.5 Funding

MF, LVB, SSW, and TSC was supported by the National Institutes of Health (NIH) National Institute of Aging (NIA) grant K08AG065519-01A1 and the Fineberg Foundation. KV was supported by NIH grants R01 NS033310, R01 AG058820, R01 AG075955, and R56 AG074473. BP was supported by NIH grants R01HG009120, R01MH115676, and R01HG006399.

### 7.6 Author Contributions

MF, BP, KV and TSC contributed to conception and design of the study. MF, LVB, and SSW performed the statistical analysis. MF wrote the first draft of the manuscript. All authors contributed to manuscript revision, read, and approved the submitted version.

## 7.7 Acknowledgments

We gratefully acknowledge the resources provided by the Institute for Precision Health (IPH) and participating UCLA ATLAS Community Health Initiative patients. The UCLA ATLAS Community Health Initiative in collaboration with UCLA ATLAS Precision Health Biobank, is a program of IPH, which directs and supports the biobanking and genotyping of biospecimen samples from participating UCLA patients in collaboration with the David Geffen School of Medicine, UCLA CTSI and UCLA Health. We would also like to acknowledge all participants and researchers at the All of Us program. The All of Us Research Program is supported by the National Institutes of Health, Office of the Director: Regional Medical Centers: 1 OT2 OD026549; 1 OT2 OD026554; 1 OT2 OD026557; 1 OT2 OD026556; 1 OT2 OD026550; 1 OT2 OD 026552; 1 OT2 OD026553; 1 OT2 OD026548; 1 OT2 OD026551; 1 OT2 OD026555; IAA #: AOD 16037; Federally Qualified Health Centers: HHSN 263201600085U; Data and Research Center: 5 U2C OD023196; Biobank: 1 U24 OD023121; The Participant Center: U24 OD023176; Participant Technology Systems Center: 1 U24 OD023163; Communications and Engagement: 3 OT2 OD023205; 3 OT2 OD023206; and Community Partners: 1 OT2 OD025277; 3 OT2 OD025315; 1 OT2 OD025337; 1 OT2 OD025276.

## Notes

### Competing Interest Statement

The authors have declared no competing interest.

### Author Declarations

As all genetic data and EHRs utilized in this study were de-identified, the study was deemed exempt from human subject research regulations (UCLA IRB# 21-000435).

