## Supplementary files for "Improving genetic risk modeling of dementia from real-world data in underrepresented populations"

**Supplementary Table 1.** GWAS information and summary results of FUMA by phenotype

| GWAS information | | | | # SNPs (overlapped with UCLA and AOU data[b]) | | |
|---|---|---|---|---|---|---|
| **Phenotype** | **Summary statistics** | **1000G reference[a]** | **N case/control** | **Candidate** | **Independent P-significant** | **Independent gene-annotated** |
| Alzheimer's Disease | Kunkle et al. (2019) | EUR | 21,982/41,944 | 1744 | 76 | 75 |
| | Kunkle et al. (2021) | AFR | 2,784/5,222 | 80 | 11 | 11 |
| | Jun et al. (2017) | ALL | 15,579/17,690 | 760 | 54 | 54 |
| Parkinson's Disease | Nalls et al. (2019) | EUR | Meta-analysis | 2640 | 27 | 27 |
| Progressive Supranuclear Palsy | Chen et al. (2018) | EUR | 1,646/10,662 | 732 | 21 | 21 |
| Lewy Body Dementia | Chia et al. (2021) | EUR | 2,981/4,391 | 356 | 9 | 9 |
| Stroke | Malik et al. (2018) | ALL | 67,162/454,450 | 903 | 21 | 20 |

*Abbreviations: AFR, African American; EUR, European; GWAS, genome-wide association study; SNP, Single-Nucleotide Polymorphism.*

**Notes**: [a] Reference panel population used for regional linkage disequilibrium patterns in identifying independent SNPs and lead P-significant SNPs. [b] Numbers of features used in PRSs building, feature selection for modeling, and phenotype prediction in the following steps.

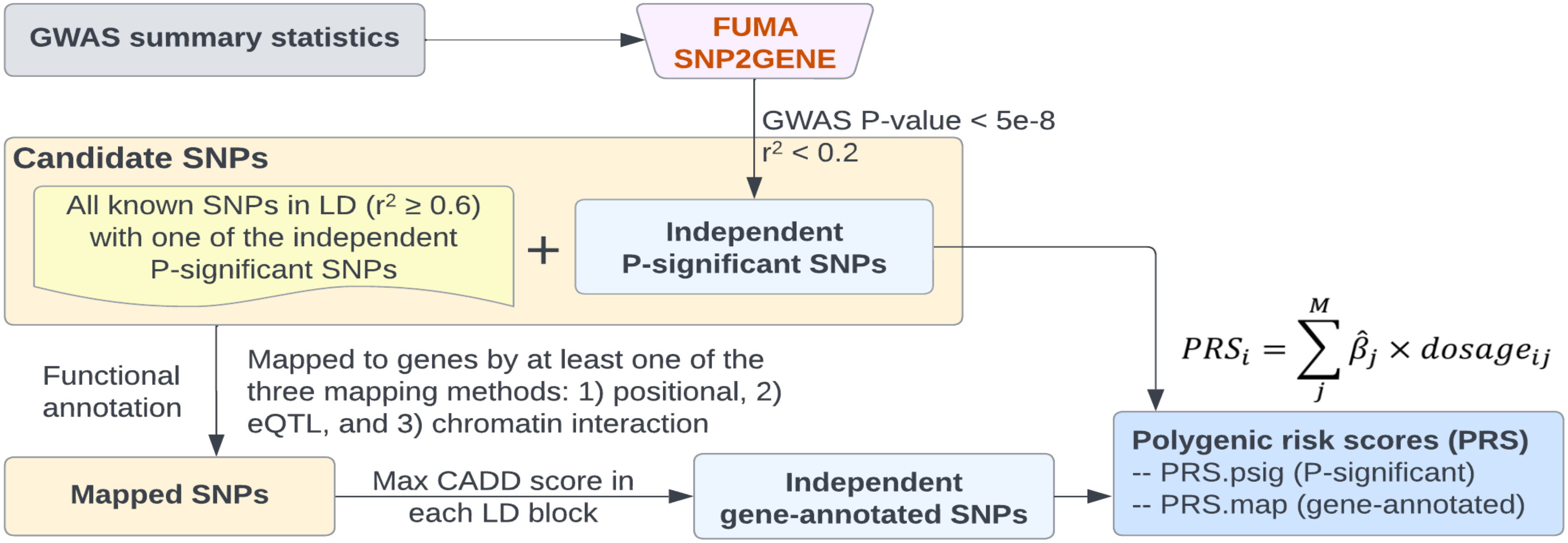


**Supplementary Figure 1. Workflow of candidate SNP selection.** Functional annotation and prioritization of SNPs using FUMA. Two distinct sets of SNPs (independent genome-wide-significant SNPs and independent gene-annotated SNPs) were identified by FUMA and subsequently used in our polygenic risk scores building and modeling steps. *Abbreviations: CADD, Combined Annotation Dependent Depletion; GWAS, genome-wide association study; LD, linkage disequilibrium; PRS, polygenic risk score; SNP, Single-Nucleotide Polymorphism.*

**Supplementary Table 2.** ICD-10 codes used for dementia phenotype definiiton

| ICD-10 code | Description |
|---|---|
| **F01** | **Vascular dementia** |
| F01.5 | Multi-infarct dementia |
| F01.50 | Multi-infarct dementia, unspecified |
| F01.51 | Multi-infarct dementia with delirium |
| F01.511 | Multi-infarct dementia with delirium in Alzheimer's disease |
| F01.518 | Multi-infarct dementia with delirium, not elsewhere classified |
| **F02.8*** | **Dementia in other diseases classified elsewhere** |
| F02.80 | Dementia in other diseases classified elsewhere, without behavioral disturbance |
| F02.81 | Dementia in other diseases classified elsewhere, with behavioral disturbance |
| F02.811 | Dementia in other diseases classified elsewhere, with behavioral disturbance in Alzheimer's disease |
| F02.818 | Dementia in other diseases classified elsewhere, with behavioral disturbance, not elsewhere classified |
| **F03** | **Unspecified dementia** |
| F03.9 | Unspecified dementia without behavioral disturbance |
| F03.90 | Unspecified dementia with delirium |
| F03.91 | Unspecified dementia with delirium in Alzheimer's disease |
| F03.911 | Unspecified dementia with delirium in Alzheimer's disease, with behavioral disturbance |
| F03.918 | Unspecified dementia with delirium in Alzheimer's disease, not elsewhere classified |
| **G30** | **Alzheimer's disease** |
| G30.0 | Alzheimer's disease with early onset |
| G30.1 | Alzheimer's disease with late onset |
| G30.8 | Other Alzheimer's disease |
| G30.9 | Alzheimer's disease, unspecified |
| **G31** | **Other degenerative diseases of nervous system, not elsewhere classified** |
| G31.0 | Pick's disease |
| G31.01 | Pick's disease with early onset |
| G31.09 | Pick's disease with late onset |
| G31.1 | Frontotemporal dementia |
| G31.83 | Dementia with Lewy bodies |
| G31.85 | Corticobasal degeneration |
| G23.1 | Parkinson's disease with dementia |

**Supplementary Table 3.** Model performance of *APOE-e4* count, polygenic risk score, and Elastic Net SNP models in dementia genetic prediction, UCLA ATLAS sample, stratified by genetic inferred ancestry[a]

| | | AUPRC | AUROC | F1 score | Accuracy | Precision | Recall | Specificity |
|---|---|---|---|---|---|---|---|---|
| **Hispanic Latino Americans (N = 610)** | | | | | | | | |
| **APOE** | e4 count | 0.373 (0.347, 0.402) | 0.652 (0.634, 0.67) | 0.454 (0.421, 0.478) | 0.673 (0.595, 0.714) | 0.394 (0.346, 0.432) | 0.547 (0.428, 0.706) | 0.714 (0.558, 0.802) |
| **AD-PRS models** | | | | | | | | |
| **AD EUR PRS** | P-significant | 0.359 (0.334, 0.389) | 0.618 (0.598, 0.641) | 0.352 (0.181, 0.457) | 0.659 (0.421, 0.764) | 0.436 (0.292, 0.684) | 0.429 (0.103, 0.929) | 0.735 (0.254, 0.984) |
| | Gene-annotated | 0.349 (0.325, 0.382) | 0.609 (0.585, 0.633) | 0.322 (0.118, 0.45) | 0.682 (0.437, 0.764) | 0.457 (0.293, 0.818) | 0.355 (0.063, 0.889) | **0.791 (0.288, 0.995)** |
| **AD AFR PRS** | P-significant | 0.374 (0.35, 0.403) | 0.645 (0.63, 0.663) | 0.438 (0.232, 0.473) | 0.658 (0.538, 0.766) | 0.39 (0.324, 0.652) | 0.548 (0.143, 0.77) | 0.694 (0.468, 0.974) |
| | Gene-annotated | 0.371 (0.345, 0.4) | 0.648 (0.631, 0.666) | 0.459 (0.433, 0.481) | 0.667 (0.597, 0.71) | 0.39 (0.347, 0.428) | 0.565 (0.46, 0.706) | 0.701 (0.561, 0.788) |
| **AD multi-ancestry PRS** | P-significant | 0.361 (0.336, 0.392) | 0.628 (0.609, 0.648) | 0.373 (0.132, 0.461) | 0.616 (0.425, 0.766) | 0.419 (0.293, 0.818) | 0.535 (0.071, 0.929) | 0.643 (0.264, 0.995) |
| | Gene-annotated | 0.363 (0.338, 0.394) | 0.641 (0.624, 0.66) | 0.444 (0.207, 0.473) | 0.521 (0.423, 0.764) | 0.338 (0.294, 0.667) | **0.792 (0.119, 0.944)** | 0.431 (0.246, 0.979) |
| **Multi-PRS models** | | | | | | | | |
| **PRSs using AD GWASs only[b]** | P-significant | 0.367 (0.34, 0.398) | 0.634 (0.612, 0.656) | 0.427 (0.212, 0.471) | 0.659 (0.559, 0.762) | 0.389 (0.327, 0.633) | 0.525 (0.127, 0.722) | 0.703 (0.51, 0.974) |
| | Gene-annotated | 0.364 (0.335, 0.395) | 0.637 (0.613, 0.658) | 0.42 (0.168, 0.467) | 0.648 (0.446, 0.766) | 0.396 (0.301, 0.769) | 0.537 (0.095, 0.913) | 0.685 (0.291, 0.992) |
| **PRSs using AD + Neuro GWASs[c]** | P-significant | 0.344 (0.31, 0.38) | 0.608 (0.575, 0.637) | 0.39 (0.144, 0.456) | 0.659 (0.532, 0.76) | 0.387 (0.31, 0.636) | 0.463 (0.079, 0.762) | 0.724 (0.46, 0.984) |
| | Gene-annotated | 0.357 (0.324, 0.393) | 0.607 (0.578, 0.634) | 0.379 (0.119, 0.462) | 0.67 (0.557, 0.766) | 0.431 (0.32, 0.889) | 0.447 (0.063, 0.699) | 0.744 (0.518, 0.997) |
| **Elastic Net SNPs models** | | | | | | | | |
| **SNPs from AD GWASs only** | P-significant | 0.398 (0.364, 0.441) | 0.68 (0.649, 0.709) | 0.476 (0.406, 0.513) | 0.617 (0.49, 0.734) | 0.374 (0.316, 0.46) | 0.706 (0.373, 0.921) | 0.587 (0.354, 0.854) |
| | Gene-annotated | 0.41 (0.378, 0.449) | 0.681 (0.651, 0.702) | 0.45 (0.296, 0.5) | 0.675 (0.53, 0.772) | 0.426 (0.329, 0.645) | 0.555 (0.19, 0.849) | 0.715 (0.423, 0.963) |
| **SNPs from AD + Neuro GWASs** | P-significant | 0.415 (0.376, 0.46) | 0.699 (0.664, 0.727) | 0.492 (0.437, 0.527) | 0.635 (0.508, 0.74) | 0.388 (0.323, 0.477) | 0.712 (0.412, 0.913) | 0.609 (0.378, 0.841) |
| | Gene-annotated | **0.451 (0.402, 0.495)** | **0.715 (0.662, 0.739)** | **0.498 (0.383, 0.541)** | **0.701 (0.597, 0.776)** | **0.447 (0.363, 0.604)** | 0.604 (0.286, 0.833) | 0.733 (0.521, 0.934) |
| **African Americans (N = 440)** | | | | | | | | |
| **APOE** | e4 count | 0.307 (0.282, 0.343) | 0.596 (0.569, 0.623) | 0.441 (0.398, 0.474) | 0.589 (0.476, 0.679) | 0.346 (0.301, 0.374) | 0.66 (0.428, 0.857) | 0.566 (0.353, 0.762) |
| **AD-PRS models** | | | | | | | | |
| **AD EUR PRS** | P-significant | 0.272 (0.249, 0.302) | 0.561 (0.525, 0.595) | 0.411 (0.367, 0.441) | 0.552 (0.327, 0.643) | 0.316 (0.266, 0.347) | 0.636 (0.428, 0.976) | 0.524 (0.111, 0.722) |
| | Gene-annotated | 0.273 (0.243, 0.314) | 0.548 (0.505, 0.584) | 0.393 (0.069, 0.44) | 0.494 (0.354, 0.753) | 0.311 (0.214, 0.6) | 0.7 (0.048, 0.917) | 0.426 (0.162, 0.988) |
| **AD AFR PRS** | P-significant | 0.277 (0.25, 0.31) | 0.566 (0.527, 0.599) | 0.402 (0.019, 0.444) | 0.492 (0.241, 0.696) | 0.293 (0.053, 0.373) | 0.715 (0.012, 0.964) | 0.418 (0, 0.917) |
| | Gene-annotated | 0.274 (0.25, 0.306) | 0.562 (0.527, 0.595) | 0.413 (0.381, 0.444) | 0.47 (0.238, 0.631) | 0.295 (0.239, 0.341) | 0.74 (0.488, 0.964) | 0.38 (0, 0.671) |
| **AD multi-ancestry PRS** | P-significant | 0.281 (0.255, 0.314) | 0.574 (0.542, 0.604) | 0.434 (0.391, 0.453) | 0.482 (0.399, 0.661) | 0.304 (0.282, 0.356) | 0.799 (0.44, 0.929) | 0.376 (0.222, 0.738) |
| | Gene-annotated | 0.282 (0.258, 0.313) | 0.581 (0.549, 0.61) | 0.44 (0.421, 0.458) | 0.477 (0.363, 0.595) | 0.302 (0.277, 0.327) | **0.823 (0.607, 0.964)** | 0.362 (0.171, 0.591) |
| **Multi-PRS models** | | | | | | | | |
| **PRSs using AD GWASs only[b]** | P-significant | 0.263 (0.235, 0.292) | 0.549 (0.502, 0.591) | 0.381 (0.02, 0.449) | 0.525 (0.39, 0.697) | 0.282 (0.062, 0.342) | 0.648 (0.012, 0.893) | 0.484 (0.226, 0.925) |
| | Gene-annotated | 0.267 (0.24, 0.298) | 0.551 (0.509, 0.589) | 0.418 (0.108, 0.453) | 0.482 (0.363, 0.634) | 0.299 (0.248, 0.324) | 0.767 (0.095, 0.941) | 0.387 (0.175, 0.81) |
| **PRSs using AD + Neuro GWASs[c]** | P-significant | 0.234 (0.209, 0.262) | 0.486 (0.428, 0.54) | 0.177 (0.018, 0.416) | 0.558 (0.244, 0.711) | 0.17 (0.04, 0.325) | 0.305 (0.012, 1) | 0.642 (0.012, 0.944) |
| | Gene-annotated | 0.252 (0.225, 0.287) | 0.508 (0.463, 0.554) | 0.291 (0.02, 0.42) | 0.454 (0.244, 0.756) | 0.288 (0.071, 1) | 0.596 (0.012, 0.988) | 0.407 (0, 1) |
| **Elastic Net SNPs models** | | | | | | | | |
| **SNPs from AD GWASs only** | P-significant | 0.395 (0.348, 0.445) | 0.655 (0.625, 0.685) | 0.424 (0.191, 0.493) | 0.677 (0.491, 0.774) | 0.442 (0.314, 0.819) | 0.509 (0.107, 0.881) | 0.733 (0.365, 0.992) |
| | Gene-annotated | 0.42 (0.369, 0.473) | 0.663 (0.632, 0.694) | 0.382 (0.154, 0.495) | 0.718 (0.5, 0.783) | **0.548 (0.319, 1)** | 0.395 (0.083, 0.893) | **0.826 (0.373, 1)** |
| **SNPs from AD + Neuro GWASs** | P-significant | 0.445 (0.389, 0.507) | 0.689 (0.655, 0.724) | 0.416 (0.252, 0.52) | **0.721 (0.503, 0.789)** | 0.54 (0.324, 0.867) | 0.436 (0.154, 0.917) | 0.816 (0.365, 0.992) |
| | Gene-annotated | **0.451 (0.397, 0.506)** | **0.698 (0.665, 0.73)** | **0.459 (0.28, 0.532)** | 0.72 (0.562, 0.789) | 0.5 (0.346, 0.827) | 0.499 (0.167, 0.845) | 0.794 (0.464, 0.988) |

*Abbreviations: AD, Alzheimer's Disease; APOE, apolipoprotein E; AUROC, Area Under the ROC Curve; AUPRC, Area Under the Precision-Recall Curve; EUR, European; GWAS, Genome-Wide Association Study; PRS, Polygenic Risk Score; SNP, Single-Nucleotide Polymorphism.*

**Notes:**
[a] All models (if not other specified) have regressed out age, sex, and ancestry-specific principal components. Thresholds were determined by maximizing absolute Matthews correlation coefficient.
[b] All AD PRSs built with EUR, AFR, and multi-ancestry GWASs using P-significant/gene-annotated SNPs were included in the model at the same time.
[c] All AD PRSs built with EUR, AFR, and multi-ancestry GWASs, and neurodegenerative disease PRS (Parkinson's disease, progressive supranuclear palsy, Lewy body dementia, and stroke) using P-significant/gene-annotated SNPs were included in the model at the same time.

**Supplementary Table 4.** Performance of models using lead SNPs ($r^2$ cut-off <0.1 for defining independent genome-wide-significant SNPs) in dementia genetic prediction, UCLA ATLAS sample, stratified by genetic inferred ancestry[a]

| | Overall performace | | Threshold maximize absolute MCC | | | | |
|---|---|---|---|---|---|---|---|
| **Hispanic Latino American ancestry sample (N = 610)** | | | | | | | |
| | **AUPRC** | **AUROC** | **F1 score** | **Accuracy** | **Precision** | **Recall** | **Specificity** |
| **AD EUR PRS** | 0.343 (0.316, 0.372) | 0.598 (0.57, 0.624) | 0.289 (0.103, 0.444) | 0.702 (0.456, 0.766) | **0.497 (0.295, 0.875)** | 0.29 (0.056, 0.841) | 0.84 (0.323, 0.997) |
| **AD AFR PRS** | 0.377 (0.35, 0.407) | 0.649 (0.631, 0.666) | 0.453 (0.414, 0.48) | 0.675 (0.567, 0.718) | 0.397 (0.337, 0.439) | 0.542 (0.413, 0.754) | 0.719 (0.51, 0.812) |
| **AD multi-ancestry PRS** | 0.369 (0.346, 0.398) | 0.624 (0.607, 0.643) | 0.342 (0.144, 0.446) | **0.71 (0.551, 0.768)** | 0.484 (0.325, 0.819) | 0.337 (0.079, 0.738) | **0.835 (0.494, 0.995)** |
| **PRSs using AD GWASs only[b]** | 0.363 (0.331, 0.396) | 0.623 (0.596, 0.65) | 0.436 (0.391, 0.467) | 0.682 (0.617, 0.724) | 0.399 (0.351, 0.443) | 0.497 (0.373, 0.643) | 0.743 (0.611, 0.844) |
| **PRSs using AD + Neuro GWASs[c]** | 0.346 (0.31, 0.384) | 0.603 (0.569, 0.635) | 0.385 (0.2, 0.446) | 0.693 (0.589, 0.762) | 0.409 (0.332, 0.632) | 0.399 (0.119, 0.651) | 0.791 (0.577, 0.976) |
| **SNPs from AD GWASs only** | 0.406 (0.371, 0.438) | 0.683 (0.648, 0.705) | 0.472 (0.387, 0.504) | 0.622 (0.486, 0.74) | 0.382 (0.316, 0.475) | **0.687 (0.341, 0.929)** | 0.6 (0.341, 0.868) |
| **SNPs from AD + Neuro GWASs** | **0.411 (0.373, 0.446)** | **0.689 (0.651, 0.712)** | **0.475 (0.398, 0.509)** | 0.628 (0.494, 0.75) | 0.386 (0.321, 0.5) | 0.683 (0.341, 0.929) | 0.61 (0.352, 0.881) |
| **African American ancestry sample (N = 440)** | | | | | | | |
| | **Overall performace** | | **Threshold maximize absolute MCC** | | | | |
| | **AUPRC** | **AUROC** | **F1 score** | **Accuracy** | **Precision** | **Recall** | **Specificity** |
| **AD EUR PRS** | 0.251 (0.214, 0.303) | 0.491 (0.428, 0.549) | 0.268 (0.02, 0.415) | 0.475 (0.241, 0.759) | 0.347 (0.067, 1) | 0.519 (0.012, 0.976) | 0.461 (0, 1) |
| **AD AFR PRS** | 0.3 (0.271, 0.34) | 0.578 (0.552, 0.606) | 0.393 (0.286, 0.433) | 0.611 (0.333, 0.741) | 0.347 (0.269, 0.455) | 0.521 (0.214, 0.976) | 0.641 (0.119, 0.917) |
| **AD multi-ancestry PRS** | 0.305 (0.277, 0.34) | 0.595 (0.57, 0.62) | 0.438 (0.421, 0.459) | 0.508 (0.408, 0.595) | 0.315 (0.285, 0.337) | **0.776 (0.619, 0.929)** | 0.418 (0.234, 0.583) |
| **PRSs using AD GWASs only[b]** | 0.315 (0.283, 0.355) | 0.582 (0.551, 0.613) | 0.367 (0.145, 0.45) | 0.576 (0.396, 0.759) | 0.373 (0.279, 0.751) | 0.568 (0.083, 0.917) | 0.578 (0.226, 0.992) |
| **PRSs using AD + Neuro GWASs[c]** | 0.3 (0.267, 0.34) | 0.575 (0.539, 0.61) | 0.404 (0.11, 0.448) | 0.508 (0.351, 0.759) | 0.333 (0.27, 0.801) | 0.713 (0.06, 0.964) | 0.439 (0.151, 0.996) |
| **SNPs from AD GWASs only** | 0.337 (0.295, 0.382) | 0.627 (0.589, 0.662) | 0.451 (0.376, 0.49) | 0.61 (0.488, 0.729) | 0.359 (0.306, 0.456) | 0.649 (0.333, 0.845) | 0.597 (0.373, 0.857) |
| **SNPs from AD + Neuro GWASs** | **0.379 (0.332, 0.429)** | **0.666 (0.634, 0.696)** | **0.476 (0.405, 0.52)** | **0.67 (0.554, 0.762)** | **0.41 (0.335, 0.533)** | 0.606 (0.345, 0.833) | **0.691 (0.464, 0.901)** |

*Abbreviations: AD, Alzheimer's Disease; AFR, African American; AUROC, Area Under the ROC Curve; AUPRC, Area Under the Precision-Recall Curve; EUR, European; GWAS, Genome-Wide Association Study; MCC, Matthews Correlation Coefficient; PRS, Polygenic Risk Score; SNP, Single-Nucleotide Polymorphism.*

**Notes:**

[a] All models (if not other specified) adjusted for age, sex, and first four ancestry-specific principal components.

[b] All AD PRSs built with EUR, AFR, and multi-ancestry GWASs using P-significant/gene-annotated SNPs were included in the model at the same time.

[c] All AD PRSs built with EUR, AFR, and multi-ancestry GWASs, and neurodegenerative disease PRS (Parkinson's disease, progressive supranuclear palsy, Lewy body dementia, and stroke) using P-significant/gene-annotated SNPs were included in the model at the same time.

**Supplementary Table 5.** Mapped genes of selected risk SNPs from the best-performing Elastic Net SNP model, UCLA ATLAS sample, by genetic inferred ancestry

| No | Gene | Symbol | CHR | Start | End | Type | posMap | eqtlMap (Direction) | ciMap |
|---|---|---|---|---|---|---|---|---|---|
| **Hispanic Latino Americans (HLA)** | | | | | | | | | |
| **No** | **Gene** | **Symbol** | **CHR** | **Start** | **End** | **Type** | **posMap** | **eqtlMap (Direction)** | **ciMap** |
| 1 | ENSG00000144659 | *SLC25A38* | 3 | 39424839 | 39438842 | protein_coding | No | Yes (+) | No |
| 2 | ENSG00000168028 | *RPSA* | 3 | 39448180 | 39454033 | protein_coding | Yes | Yes (+) | No |
| 3 | ENSG00000185619 | *PCGF3* | 4 | 699537 | 764428 | protein_coding | Yes | Yes (-) | No |
| 4 | ENSG00000249519 | *RP11-777N19.1* | 4 | 111715559 | 111718500 | lincRNA | Yes | No | No |
| 5 | ENSG00000137642 | *SORL1* | 11 | 121322912 | 121504402 | protein_coding | Yes | No | No |
| 6 | ENSG00000266903 | *CTB-171A8.1* | 19 | 45135500 | 45222031 | antisense | Yes | No | No |
| 7 | ENSG00000252200 | *snoZ6* | 19 | 45229248 | 45229322 | snoRNA | Yes | No | No |
| 8 | ENSG00000187244 | *BCAM* | 19 | 45312328 | 45324673 | protein_coding | Yes | No | No |
| 9 | ENSG00000130202 | *PVRL2* | 19 | 45349432 | 45392485 | protein_coding | Yes | No | No |
| 10 | ENSG00000130204 | *TOMM40* | 19 | 45393826 | 45406946 | protein_coding | Yes | No | No |
| 11 | ENSG00000130203 | *APOE* | 19 | 45409011 | 45412650 | protein_coding | Yes | No | No |
| 12 | ENSG00000130208 | *APOC1* | 19 | 45417504 | 45422606 | protein_coding | Yes | No | No |
| 13 | ENSG00000214855 | *APOC1P1* | 19 | 45430061 | 45434643 | pseudogene | No | No | Yes |
| 14 | ENSG00000104859 | *CLASRP* | 19 | 45542298 | 45574214 | protein_coding | No | No | Yes |
| **African Americans (AA)** | | | | | | | | | |
| **No** | **Gene** | **Symbol** | **CHR** | **Start** | **End** | **Type** | **posMap** | **eqtlMap (Direction)** | **ciMap** |
| 1 | ENSG00000136717 | *BIN1* | 2 | 127805603 | 127864931 | protein_coding | Yes | No | No |
| 2 | ENSG00000159840 | *ZYX* | 7 | 143078173 | 143088204 | protein_coding | Yes | No | No |
| 3 | ENSG00000050327 | *ARHGEF5* | 7 | 144052381 | 144077725 | protein_coding | No | Yes (+) | No |
| 4 | ENSG00000080854 | *IGSF9B* | 11 | 133778459 | 133826880 | protein_coding | Yes | No | No |
| 5 | ENSG00000255406 | *RP11-713P17.5* | 11 | 133896882 | 133898013 | lincRNA | No | No | Yes |
| 6 | ENSG00000166086 | *JAM3* | 11 | 133938820 | 134021896 | protein_coding | No | No | Yes |
| 7 | ENSG00000254481 | *PTP4A2P2* | 11 | 133993723 | 133994224 | pseudogene | No | No | Yes |
| 8 | ENSG00000180329 | *CCDC43* | 17 | 42750437 | 42767147 | protein_coding | No | No | Yes |
| 9 | ENSG00000161692 | *DBF4B* | 17 | 42785976 | 42829632 | protein_coding | No | Yes (-) | No |
| 10 | ENSG00000073670 | *ADAM11* | 17 | 42836399 | 42859214 | protein_coding | No | No | Yes |
| 11 | ENSG00000131095 | *GFAP* | 17 | 42982376 | 42994305 | protein_coding | No | No | Yes |
| 12 | ENSG00000131094 | *C1QL1* | 17 | 43037061 | 43045439 | protein_coding | Yes | Yes (+) | No |
| 13 | ENSG00000267788 | *CTD-2534I21.9* | 17 | 43059882 | 43060140 | lincRNA | No | No | Yes |
| 14 | ENSG00000267282 | *CTB-129P6.4* | 19 | 45385284 | 45394133 | antisense | Yes | No | No |
| 15 | ENSG00000130204 | *TOMM40* | 19 | 45393826 | 45406946 | protein_coding | Yes | No | No |
| 16 | ENSG00000130203 | *APOE* | 19 | 45409011 | 45412650 | protein_coding | Yes | Yes (-) | No |
| 17 | ENSG00000214855 | *APOC1P1* | 19 | 45430061 | 45434643 | pseudogene | Yes | Yes (+) | Yes |
| 18 | ENSG00000104859 | *CLASRP* | 19 | 45542298 | 45574214 | protein_coding | No | No | Yes |

**Supplementary Table 6A.** Descriptive statistics of demographic and electronic health record features by case/control groups, UCLA ATLAS sample (East Asian American ancestry group, N = 673)

| | **Cases** | **Controls** | **P value** |
|---|---|---|---|
| N | 75 | 598 | - |
| Age | 80.1 (73.2, 83.5) | 76.3 (72.9, 80.7) | 0.04* |
| Sex (Female) | 48 (64%) | 325 (54%) | 0.11 |
| Span of records (in yrs) | 5.3 (2.7, 7.8) | 9.8 (8.0, 12.0) | <0.001* |
| Encounters per year | 17 (9, 28) | 12 (7, 18) | 0.002* |
| Number of encounters | 73 (23, 137) | 121 (68, 185) | <0.001* |
| Number of unique diagnosis | 64 (35, 92) | 60 (38, 90) | 0.80 |

*Abbreviations: EHR, electronic health record.*

**Notes**: Continuous variables were reported as median (IQR), and categorical variables were reported as n (%). P-values were calcualted based on Wilcoxon rank sum test or Pearson's Chi-squared test as appropriate. * Statistical siginificant at level 0.05.

**Supplementary Table 6B.** Model performance of *APOE-e4* count, polygenic risk score, and Elastic Net SNP models in dementia genetic prediction, UCLA ATLAS sample (East Asian American ancestry group, N = 673)[a]

| | | AUPRC | AUROC | F1 score | Accuracy | Precision | Recall | Specificity |
|---|---|---|---|---|---|---|---|---|
| **APOE** | e4 count | 0.463 (0.409, 0.516) | 0.708 (0.678, 0.733) | 0.476 (0.28, 0.539) | 0.725 (0.593, 0.793) | 0.502 (0.361, 0.818) | 0.523 (0.173, 0.827) | 0.792 (0.52, 0.987) |
| | | **AD-PRS models** | | | | | | |
| **AD EUR PRS** | P-significant | 0.442 (0.396, 0.493) | 0.688 (0.658, 0.715) | 0.425 (0.23, 0.516) | 0.712 (0.513, 0.787) | 0.524 (0.328, 0.9) | 0.468 (0.133, 0.907) | 0.793 (0.382, 0.996) |
| | Gene-annotated | 0.434 (0.392, 0.482) | 0.684 (0.651, 0.714) | 0.416 (0.212, 0.513) | 0.703 (0.493, 0.787) | 0.523 (0.32, 0.909) | 0.473 (0.12, 0.92) | 0.78 (0.355, 0.996) |
| **AD AFR PRS** | P-significant | 0.457 (0.408, 0.51) | 0.703 (0.673, 0.728) | 0.469 (0.28, 0.532) | 0.723 (0.573, 0.79) | 0.503 (0.352, 0.833) | 0.512 (0.173, 0.84) | 0.794 (0.484, 0.991) |
| | Gene-annotated | 0.452 (0.404, 0.501) | 0.698 (0.669, 0.722) | 0.462 (0.264, 0.529) | 0.72 (0.583, 0.787) | 0.5 (0.355, 0.834) | 0.507 (0.16, 0.827) | 0.791 (0.502, 0.991) |
| **AD multi-ancestry PRS** | P-significant | 0.459 (0.407, 0.511) | 0.702 (0.671, 0.727) | 0.46 (0.27, 0.533) | 0.729 (0.587, 0.79) | **0.519 (0.358, 0.857)** | 0.49 (0.16, 0.813) | 0.808 (0.507, 0.991) |
| | Gene-annotated | 0.454 (0.406, 0.506) | 0.697 (0.666, 0.722) | 0.457 (0.264, 0.53) | 0.727 (0.6, 0.79) | 0.514 (0.36, 0.857) | 0.487 (0.16, 0.787) | 0.807 (0.533, 0.991) |
| | | **Multi-PRS models** | | | | | | |
| **PRSs using AD GWASs only[b]** | P-significant | 0.454 (0.401, 0.507) | 0.697 (0.667, 0.723) | 0.461 (0.267, 0.529) | 0.727 (0.6, 0.79) | 0.511 (0.358, 0.842) | 0.492 (0.16, 0.787) | **0.805 (0.542, 0.991)** |
| | Gene-annotated | 0.45 (0.4, 0.501) | 0.698 (0.665, 0.726) | 0.461 (0.253, 0.527) | 0.721 (0.563, 0.787) | 0.499 (0.346, 0.833) | 0.501 (0.147, 0.84) | 0.795 (0.471, 0.991) |
| **PRSs using AD + Neuro GWASs[c]** | P-significant | 0.453 (0.402, 0.51) | 0.699 (0.666, 0.728) | 0.459 (0.253, 0.532) | 0.722 (0.553, 0.79) | 0.508 (0.342, 0.846) | 0.5 (0.147, 0.867) | 0.796 (0.458, 0.991) |
| | Gene-annotated | 0.455 (0.404, 0.513) | 0.712 (0.676, 0.743) | 0.489 (0.323, 0.549) | 0.708 (0.57, 0.787) | 0.471 (0.35, 0.739) | 0.577 (0.213, 0.853) | 0.751 (0.48, 0.973) |
| | | **Elastic Net SNPs models** | | | | | | |
| **SNPs from AD GWASs only** | P-significant | 0.474 (0.403, 0.542) | 0.721 (0.674, 0.764) | 0.489 (0.308, 0.562) | 0.712 (0.553, 0.793) | 0.491 (0.349, 0.8) | 0.576 (0.187, 0.92) | 0.758 (0.435, 0.982) |
| | Gene-annotated | 0.481 (0.406, 0.553) | 0.73 (0.674, 0.776) | 0.505 (0.312, 0.575) | 0.711 (0.58, 0.79) | 0.481 (0.358, 0.786) | 0.615 (0.2, 0.88) | 0.743 (0.476, 0.982) |
| **SNPs from AD + Neuro GWASs** | P-significant | 0.499 (0.406, 0.582) | 0.743 (0.674, 0.795) | 0.521 (0.33, 0.599) | 0.729 (0.603, 0.8) | 0.503 (0.368, 0.734) | 0.608 (0.213, 0.88) | 0.77 (0.52, 0.973) |
| | Gene-annotated | **0.511 (0.405, 0.602)** | **0.754 (0.675, 0.808)** | **0.535 (0.343, 0.613)** | **0.734 (0.61, 0.803)** | 0.505 (0.373, 0.727) | **0.627 (0.227, 0.88)** | 0.77 (0.533, 0.969) |

*Abbreviations: AD, Alzheimer's Disease; APOE, apolipoprotein E; AUROC, Area Under the ROC Curve; AUPRC, Area Under the Precision-Recall Curve; EUR, European; GWAS, Genome-Wide Association Study; PRS, Polygenic Risk Score; SNP, Single-Nucleotide Polymorphism.*

**Notes:**

[a] All models (if not other specified) have regressed out age, sex, and ancestry-specific principal components. Thresholds were determined by maximizing absolute Matthews correlation coefficient.

[b] All AD PRSs built with EUR, AFR, and multi-ancestry GWASs using P-significant/gene-annotated SNPs were included in the model at the same time.

[c] All AD PRSs built with EUR, AFR, and multi-ancestry GWASs, and neurodegenerative disease PRS (Parkinson's disease, progressive supranuclear palsy, Lewy body dementia, and stroke) using P-significant/gene-annotated SNPs were included in the model at the same time.

**Supplementary Table 6C.** Selected risk SNPs from the best-performing Elastic Net SNP model, UCLA ATLAS sample (East Asian American ancestry group, N = 673)

| rsID | CHR | POS | Variable Importance (percentage, 95% CI) | Nearest Gene | AD EUR | AD AFR | AD multi | LBD | PD | PSP | Stroke |
|---|---|---|---|---|---|---|---|---|---|---|---|
| rs429358 | 19 | 44908684 | 0.127 (0.036, 0.26) | *APOE* | | x | | | | | |
| rs35106910 | 19 | 44781009 | 0.114 (0.021, 0.298) | *CBLC* | x | | | | | | |
| rs66626994 | 19 | 44924977 | 0.076 (0.009, 0.176) | *APOC1P1* | | | x | x | | | |
| rs483082 | 19 | 44912921 | 0.067 (0.007, 0.154) | *APOC1* | | x | x | | | | |
| rs59193782 | 17 | 45357199 | 0.053 (0.003, 0.136) | *CTB-39G8.2* | | | | | | x | |
| chr1:207368589:D | 1 | 207195244 | 0.053 (0.003, 0.138) | *RP11-164O23.7* | x | | | | | | |
| rs34096562 | 8 | 16843772 | 0.053 (0.003, 0.137) | *RP11-13N12.1* | | | | | x | | |
| rs11724804 | 4 | 971991 | 0.051 (0.003, 0.14) | *DGKQ* | | | | x | x | | |
| rs6857 | 19 | 44888997 | 0.05 (0.004, 0.126) | *PVRL2* | | x | | | | | |
| rs7613 | 17 | 45394115 | 0.047 (0.003, 0.122) | *ARHGAP27* | | | | | | x | |
| rs2075650 | 19 | 44892362 | 0.045 (0.003, 0.111) | *TOMM40* | | x | x | x | | | |
| rs10769263 | 11 | 47395632 | 0.042 (0.003, 0.108) | *RP11-750H9.5* | x | | | | | | |
| rs117421612 | 17 | 45345796 | 0.029 (0.003, 0.056) | *RNA5SP443* | | | | | | x | |

*Abbreviations: AD, Alzheimer's Disease; AFR, African American; CI, confidence interval; EUR, European; LBD, Lewy body dementia; PD, Parkinson's disease; PRS, Polygenic Risk Score; PSP, progressive supranuclear palsy;SNP, Single-Nucleotide Polymorphism.*

**Supplementary Table 6D.** Mapped genes of selected risk SNPs from the best-performing Elastic Net SNP model, UCLA ATLAS sample (East Asian American ancestry group, N = 673)

| No | Gene | Symbol | CHR | Start | End | Type | posMap | eqtlMap (Direction) | ciMap |
|---|---|---|---|---|---|---|---|---|---|
| 1 | ENSG00000203710 | CR1 | 1 | 207669492 | 207813992 | protein_coding | No | Yes (+) | No |
| 2 | ENSG00000127419 | TMEM175 | 4 | 926175 | 952444 | protein_coding | No | Yes (-) | No |
| 3 | ENSG00000145214 | DGKQ | 4 | 952675 | 980683 | protein_coding | Yes | Yes (-) | No |
| 4 | ENSG00000145217 | SLC26A1 | 4 | 972861 | 987228 | protein_coding | No | Yes (+) | No |
| 5 | ENSG00000253496 | RP11-13N12.1 | 8 | 16534414 | 16772553 | lincRNA | Yes | No | No |
| 6 | ENSG00000175220 | ARHGAP1 | 11 | 46698630 | 46722165 | protein_coding | No | Yes (+) | No |
| 7 | ENSG00000255197 | RP11-750H9.5 | 11 | 47404699 | 47430741 | antisense | Yes | Yes (-) | No |
| 8 | ENSG00000165915 | SLC39A13 | 11 | 47428683 | 47438047 | protein_coding | No | Yes (-) | No |
| 9 | ENSG00000213619 | NDUFS3 | 11 | 47586888 | 47606114 | protein_coding | No | Yes (-) | No |
| 10 | ENSG00000196666 | FAM180B | 11 | 47608198 | 47610746 | protein_coding | No | Yes (-) | No |
| 11 | ENSG00000184922 | FMNL1 | 17 | 43298811 | 43324687 | protein_coding | No | Yes (+) | No |
| 12 | ENSG00000233175 | CTD-2020K17.3 | 17 | 43315395 | 43319101 | antisense | No | Yes (+) | No |
| 13 | ENSG00000006062 | MAP3K14 | 17 | 43340488 | 43394414 | processed_transcript | No | No | Yes |
| 14 | ENSG00000199953 | RNA5SP443 | 17 | 43404732 | 43404863 | rRNA | Yes | No | No |
| 15 | ENSG00000267446 | CTB-39G8.2 | 17 | 43448768 | 43449423 | lincRNA | Yes | No | No |
| 16 | ENSG00000159314 | ARHGAP27 | 17 | 43471275 | 43511787 | protein_coding | Yes | Yes (-) | No |
| 17 | ENSG00000214425 | LRRC37A4P | 17 | 43578685 | 43627701 | pseudogene | No | Yes (+) | No |
| 18 | ENSG00000263503 | RP11-707O23.5 | 17 | 43678235 | 43679706 | pseudogene | No | Yes (-) | No |
| 19 | ENSG00000204650 | CRHR1-IT1 | 17 | 43697694 | 43725582 | pseudogene | No | Yes (-) | No |
| 20 | ENSG00000185294 | SPPL2C | 17 | 43922256 | 43924438 | protein_coding | No | Yes (-) | No |
| 21 | ENSG00000262500 | RP11-259G18.2 | 17 | 44320972 | 44322410 | pseudogene | No | Yes (-) | No |
| 22 | ENSG00000261575 | RP11-259G18.1 | 17 | 44344403 | 44346060 | pseudogene | No | Yes (-) | No |
| 23 | ENSG00000142273 | CBLC | 19 | 45281126 | 45303891 | protein_coding | Yes | No | No |
| 24 | ENSG00000130204 | TOMM40 | 19 | 45393826 | 45406946 | protein_coding | Yes | No | No |
| 25 | ENSG00000130203 | APOE | 19 | 45409011 | 45412650 | protein_coding | Yes | No | No |
| 26 | ENSG00000130208 | APOC1 | 19 | 45417504 | 45422606 | protein_coding | Yes | No | No |
| 27 | ENSG00000214855 | APOC1P1 | 19 | 45430061 | 45434643 | pseudogene | Yes | No | Yes |
| 28 | ENSG00000104859 | CLASRP | 19 | 45542298 | 45574214 | protein_coding | No | No | Yes |

**Supplementary Table 7.** Descriptive statistics of demographic and electronic health record features, by data sources (All of Us vs. UCLA ATLAS, stratified by the genetic inferred group)

| | Hispanic Latino Americans | | | African Americans | | |
|---|---|---|---|---|---|---|
| **Characteristic** | **All of Us** | **UCLA ATLAS** | **p-value** | **All of Us** | **UCLA ATLAS** | **p-value** |
| N | 526 | 610 | - | 2,644 | 440 | - |
| Age | 74.0 (71.9, 78.2) | 75.8 (72.5, 80.5) | <0.001* | 70.6 (67.6, 74.9) | 75.8 (72.5, 80.5) | <0.001* |
| Sex (Female) | 331 (63%) | 372 (61%) | 0.5 | 1,735 (66%) | 264 (60%) | 0.02* |
| Span of records (in yrs) | 6.5 (5.7, 7.6) | 9.1 (6.8, 10.2) | <0.001* | 6.56 (6.08, 7.13) | 9.77 (7.27, 11.02) | <0.001* |
| Encounters per year | 9 (5, 14) | 14 (8, 22) | <0.001* | 9 (5, 15) | 13 (8, 22) | <0.001* |
| Number of encounters | 59 (32, 104) | 117 (64, 198) | <0.001* | 62 (34, 107) | 129 (71, 209) | <0.001* |
| Number of unique diagnosis | 46 (28, 69) | 71 (45, 109) | <0.001* | 44 (27, 66) | 71 (45, 103) | <0.001* |
| Dementia (Yes) | 81 (15%) | 126 (21%) | 0.02* | 181 (6.8%) | 84 (19%) | <0.001* |
| *APOE-e4* count | | | <0.001* | | | <0.001* |
| 0 | 486 (92%) | 491 (80%) | | 2,127 (80%) | 284 (65%) | |
| 1 | 38 (7.2%) | 117 (19%) | | 482 (18%) | 137 (31%) | |
| 2 | 2 (0.4%) | 2 (0.3%) | | 35 (1.3%) | 19 (4.3%) | |

**Notes**: Continuous variables were reported as median (IQR), and categorical variables were reported as n (%). P-values were calcualted based on Wilcoxon rank sum test or Pearson's Chi-squared test as appropriate. * Statistical siginificant at level 0.05.

**Supplementary Table 8.** Descriptive statistics of demographic and electronic health record features by case/control groups, All of Us sample, stratified by genetic ancestry

| | **Hispanic Latino Americans (N = 610)** | | | **African Americans (N = 2,644)** | | |
|---|---|---|---|---|---|---|
| | **Cases** | **Controls** | **P value** | **Cases** | **Controls** | **P value** |
| N | 81 | 445 | - | 181 | 2,463 | - |
| Age | 73.1 (68.4, 80.1) | 74.0 (72.0, 78.0) | 0.03* | 70.7 (65.7, 77.8) | 70.5 (67.7, 74.8) | 0.4 |
| Sex (Female) | 50 (62%) | 281 (63%) | 0.80 | 113 (62%) | 1,622 (66%) | 0.30 |
| Span of records (in yrs) | 4.4 (2.6, 6.1) | 6.6 (6.0, 7.9) | <0.001* | 4.25 (2.12, 6.14) | 6.58 (6.18, 7.25) | <0.001* |
| Encounters per year | 10 (5, 18) | 8 (5, 14) | 0.14 | 14 (7, 23) | 9 (5, 15) | <0.001* |
| Number of encounters | 36 (15, 76) | 62 (35, 107) | <0.001* | 52 (20, 96) | 63 (34, 108) | <0.001* |
| Number of unique diagnosis | 47 (25, 68) | 45 (28, 69) | 0.80 | 51 (31, 76) | 43 (27, 66) | 0.01* |

**Notes**: Continuous variables were reported as median (IQR), and categorical variables were reported as n (%). P-values were calcualted based on Wilcoxon rank sum test or Pearson's Chi-squared test as appropriate. * Statistical siginificant at level 0.05.

**Supplementary Table 9.** Pearson's correlation of polygenic risk scores that built with genome-wide-significant or gene-annotated SNPs, UCLA ATLAS sample, stratified by genetic inferred ancestry

| | **Pearson's Correlation** | |
|---|---|---|
| **Polygenic risk score GWAS** | **Hispanic Latino American** | **African American** |
| Alzheimer's Disease (European ancestry) | 0.885 | 0.837 |
| Alzheimer's Disease (African ancestry) | 0.953 | 0.963 |
| Alzheimer's Disease (multi-ancestry) | 0.953 | 0.919 |
| Lewy Body Dementia | 0.714 | 0.698 |
| Parkinson's Disease | 0.561 | 0.29 |
| Progressive Supranuclear Palsy | 0.813 | 0.657 |
| Stroke | 0.737 | 0.669 |

*Abbreviations: GWAS, genome-wide association study, SNP, Single-Nucleotide Polymorphism.*

| | |
|---|---|
| AA | African American |
| AD | Alzheimer's disease |
| APOE | Apolipoprotein E |
| AUPRC | Area Under the Precision-Recall Curve |
| AUROC | area under the receiver operating characteristic |
| CADD | Combined Annotation-Dependent Depletion |
| CI | confidence intervals |
| EA | European American |
| EAA | East Asian American |
| EHR | Electronic Health Records |
| FTD | Frontotemporal dementia |
| FUMA | Functional Mapping and Annotation of Genome-Wide Association Studies |
| GIA | Genetic Inferred Ancestry |
| GO | Gene Ontology |
| GWAS | Genome-Wide Association Studies |
| HLA | Hispanic Latino American |
| LBD | Lewy body dementia |
| LD | Linkage disequilibrium |
| MCC | Matthews Correlation Coefficient |
| PC | principal components |
| PDD | Parkinson’s disease dementia |
| PRS | Polygenic risk scores |
| SAA | South Asian American |
| SNP | Single-Nucleotide Polymorphisms |
